# Ancestry diversity in the genetic determinants of the human plasma proteome enhances identification of potential drug targets

**DOI:** 10.1101/2023.11.13.23298365

**Authors:** Alfred Pozarickij, Saredo Said, Kuang Lin, Sam Morris, Ahmed Edris, Andri Iona, Christiana Kartsonaki, Neil Wright, Pang Yao, Hannah Fry, Yiping Chen, Ling Yang, Huaidong Du, Derrick Bennett, Daniel Avery, Dan Valle Schmidt, Jun Lv, Canqing Yu, Dianjianyi Sun, Pei Pei, Junshi Chen, Michael Hill, Richard Peto, Rory Collins, Robert Clarke, Liming Li, Iona Y Millwood, Zhengming Chen, Robin G Walters, China Kadoorie Biobank Collaborative Group

## Abstract

The proteome is fundamental to human biology and health but little is known about ancestral diversity of its genetic determinants. In GWAS of plasma levels of 2,923 proteins in 3,974 Chinese adults, we identified pQTLs for 1,784 proteins, including 1,312 *cis*-pQTLs for 1,264 proteins. Fine-mapping identified 3,475 credible sets for independent pQTLs, 36% of which were distinct from those identified in European adults as assessed by three different methods. In phenome-wide MR analyses, 59 disease associations were in strong LD (r^2^>0.95) with a *cis*-pQTL sentinel variant, including 26 with strong evidence of colocalisation (PP.H4>0.9) and 34 not identified in previous similar European studies. Evaluation of current drug development identified 8 confirmed therapeutic targets, 8 potential repurposing targets, and 19 potential novel targets (13 not previously reported in Europeans). The findings demonstrate the importance of extending proteogenomic studies to non-European ancestry populations to identify potential novel drug targets for major diseases.

## Introduction

Proteins mediate the effects of genes in determining body structure and function, and play a key role in human health and disease^1,2^. Differences in the expression or activity of a wide range of proteins have been implicated in disease processes and, accordingly, most existing drug targets are proteins^3,4^. Nevertheless, while the Human Protein Atlas (version 24.0)^5^ identifies 4,906 proteins with experimental evidence of disease involvement, the large majority (82%) are yet to be evaluated as potential drug targets in experimental studies.

Identification and assessment of proteins as potential drug targets requires accurate measurement of their levels in the relevant tissue. Advances in high-throughput proteomics assays now enable measurement of levels in plasma of several thousand proteins simultaneously^6–8^, reflecting secretion into blood, cellular leakage, fragments released from membrane proteins, or exogenous proteins from infectious organisms. Thus, application of these novel assays in large clinical and population studies in diverse populations, and integrated analysis with genetic and disease outcome data, affords new research opportunities for hypothesis testing and drug target discovery^9^, as well as for improved risk prediction^10^, early detection, and diagnosis of specific diseases^11–14^. In particular, identification of the genetic determinants of protein levels^15,16^ (i.e. protein quantitative trait loci, pQTLs) provide instruments for use in Mendelian randomisation (MR) analyses, enabling assessment of the causal relevance of individual proteins for particular diseases or traits (e.g. heart disease, stroke, diabetes, adiposity)^17–19^, thereby identifying potential novel or repurposing drug targets^15^.

To date, analysis of the genetic architecture of the plasma proteome has mainly focussed on European ancestry (EUR) populations^15,20^, including the recent UK Biobank Pharma Proteomics Project (UKB-PPP)^21^ which identified pQTLs for 2923 proteins measured by the Olink Explore 3072 PEA platform^21^. However, the discovery cohort comprised individuals of European descent and replication was performed in only a small number of individuals from non-EUR populations^21^. Important examples of protein-altering variants absent or rare in EUR but common in other ancestries^22–24^ have highlighted that there are benefits from extending proteogenomic analyses to diverse populations. As well as greatly extending the range of pQTLs available for downstream analyses beyond those identified in EUR populations, additional instruments can generate allelic series with varying magnitudes of effect or tissue-specificity. In addition, differing LD patterns in diverse populations can help to reduce the likelihood of confounding by LD, and assist in identification of causal variants. These can strengthen causal inference of the role of proteins in disease outcomes, with a positive impact on the effectiveness of genomics-guided therapeutic interventions^25,26^ – genetic support predicts improved progression and approval of new drug developments by more than 2-fold^27^, with two-thirds of new FDA approved drugs in 2021 supported by genetic evidence^28^.

Although some attempts have been made to discover pQTLs in non-EUR ancestries, little is known about the extent to which ancestry diversity can contribute to the range of pQTLs available for analysis^15,20^. In a recent Japanese study of the Olink Explore platform among 1,384 COVID patients, many of the associated variants were absent from the UKB-PPP dataset so that their replication could not readily be assessed^29^. Likewise, using a mass-spectrometry platform not widely used in other studies, a study in 2,958 Han Chinese assayed 304 unique proteins which have little overlap with available data on other platforms^30^. Nevertheless, a study of 4,657 proteins assayed on the SomaScan platform which included 1,871 African Americans (AA) showed that one third of AA lead variants were rare or absent in EUR^31^, but the much greater genetic diversity in African populations may not be reflected in other global populations. Both these latter studies focussed primarily on *cis*-pQTLs, so that there is even more limited information on *trans*-pQTLs in non-EUR populations.

To address the need for more comprehensive analyses of the genetic architecture of the plasma proteome in non-EUR populations, we report GWAS analyses of 2,923 proteins measured on the Olink Explore 3072 PEA platform in ∼4,000 adults from the China Kadoorie Biobank (CKB). We also present a detailed comparison of the proteogenomic architecture of EAS and EUR populations. Moreover, we further assess the extent to which ancestry diversity can enhance investigation of the causal relevance of plasma proteins and of their potential as drug targets for different diseases.

## Results

### pQTL discovery

Figure 1 summarises the study design, analytic approaches, and main findings. The characteristics of the 3,977 unrelated individuals included in the study are provided in **Supplementary Table 1**. The 2,923 proteins were assayed in two batches, with 39 having visible multi-modal distributions, of which 35 also had multi-modal distributions in the previous UKB-PPP study (**Supplementary** Figure 1**, Supplementary Table 2**). GWAS analyses (in 3,974 or 3,968 individuals for batch 1 [1,463 proteins] or batch 2 [1,460 proteins], respectively) (**Supplementary Table 3**) identified 3,196 pQTLs at genome-wide significance (P≤5x10^-8^) for 1,784 proteins, with 1,264 proteins (43%) having at least one *cis*-pQTL (defined as those with a lead variant within 1Mbp of the corresponding protein structural gene) (Figure 2A, **Supplementary Table 4**). Although only 57% of these pQTLs (1,819 for 1,313 proteins) passed the more stringent global Bonferroni-adjusted significance threshold used by UKB-PPP (i.e. P<1.71x10^-11^ = 5x10^-8^/2,923), this mostly reflected weaker associations for *trans*-pQTLs, with only 37% (691 of 1,884) passing this threshold, compared with 86% (1,128 out of 1,312) of *cis* associations.

**Figure 1.**
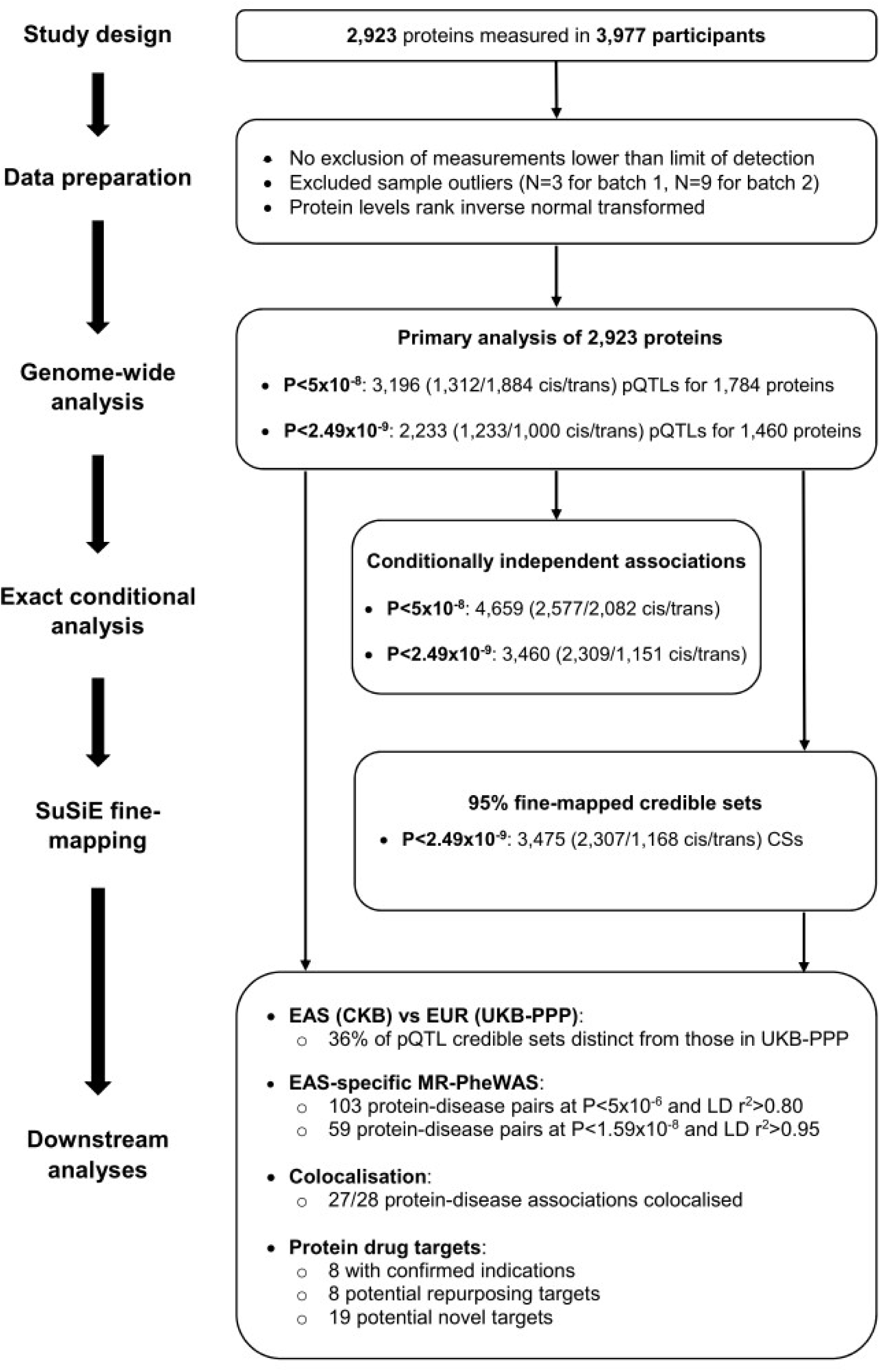
Summary of study design, analytic approaches. and key findings.

**Figure 2.**
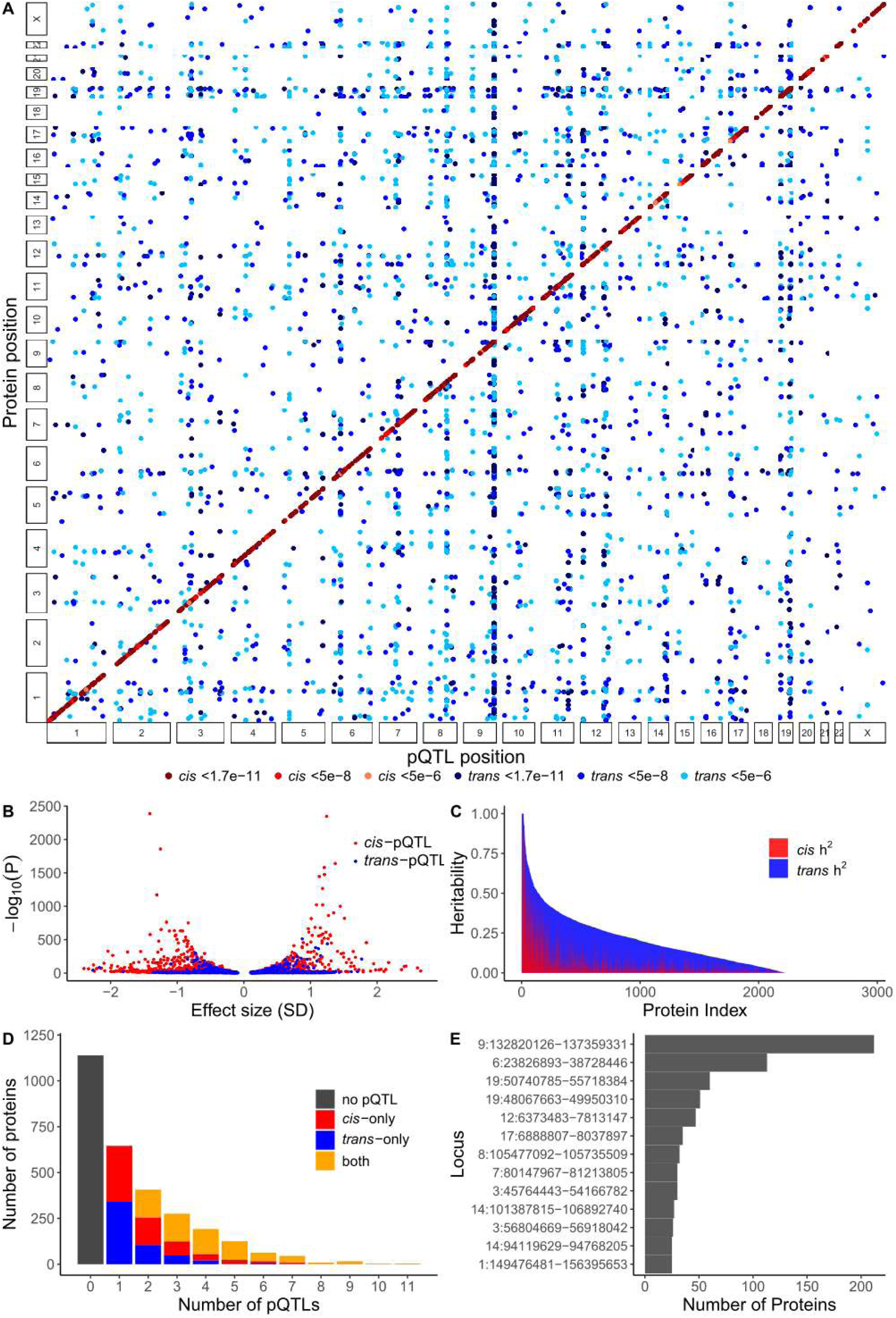
The genetic architecture of East Asian pQTLs. **(A)** Genomic positions of all conditionally independent pQTLs at P<5x10^-8^ in relation to the locations of the structural genes for associated proteins. Colour depth reflects strength of association, and includes suggestive associations at P<5x10^-6^. **(B)** pQTL effect estimates. Relationship between strength of association and minor allele effect size for all conditionally-independent association signals. **(C)** Heritability of transformed protein levels. Polygenic *cis* and *trans* heritability is shown for all proteins, ordered according to total estimated heritability. **(D)** Multiple pQTLs. Frequency distribution of proteins with different numbers of independent pQTL associations, according to whether their pQTLs are *cis* only, *trans* only, or both. **(E)** pQTL pleiotropy. Number of protein associations at a given locus, for all loci associated with at least 25 proteins at P<5x10^-8^. In all panels, *cis-* and *trans-*pQTLs are shown in red and blue, respectively.

Replication of sentinel variants or proxies for 1,174 CKB *cis*-pQTLs with available summary data from the previous GWAS of Olink-assayed proteins by the Japan COVID-19 Task Force^29^ showed consistent effect sizes with 1,168 (99.5%) nominally significant (P<0.05) and 807 (69%) reaching genome-wide significance (P<5x10^-8^) (**Supplementary** Figure 2, **Supplementary Table 5**). Despite the larger sample size in our study, we replicated a lower proportion of *cis*-pQTLs identified by the Japanese study, perhaps reflecting inclusion of loci influencing responses to COVID infection: out of 1,042 lead variants or proxies included in the CKB GWAS, 879 (84%) were associated in CKB at genome-wide significance, while 36 failed to reach nominal significance in CKB (**Supplementary** Figure 3, **Supplementary Table 6**).

Exact conditional analysis identified an additional 1,463 conditionally-independent associations at P<5x10^-8^, of which 1,265 and 198 were at *cis* and *trans* loci, respectively (**Supplementary Table 7**).

Across all 4,659 associations, conditional effect size estimates for the minor alleles of lead variants at *cis-*pQTLs were generally larger than at *trans*-pQTLs, and had directions of effect skewed towards reductions in plasma protein levels (Figure 2B): Per-allele effect estimates for *cis* associations ranged from –2.40 to +2.65 SDs (median=–0.18; mean=–0.10; mean absolute effect=0.54) and for *trans*- pQTLs effect estimates ranged from -2.26 to +1.77 (median +0.13; mean=+0.03; mean absolute effect=0.38); minor alleles were associated with lower protein levels for 56% and 47% of associated variants at *cis*- and *trans*-pQTLs, respectively.

Ppolygenic genome-wide narrow-sense heritability (h^2^), estimated after adjustment for GWAS covariates, ranged from near zero (705 proteins with h^2^ <0.1%) to virtually 100% (16 proteins with h^2^ >99.9%, including HLA-DRA, IFNGR2, and IL6R) (Figure 2C, **Supplementary Table 8**). Although 1,659 proteins (including 70 encoded on chrX) had no identified *cis*-pQTL, estimates of *cis*-heritability derived as the difference between genome-wide h^2^ estimates with and without exclusion of the *cis* region found only 1,155 autosomal proteins with near-zero *cis*-h^2^ (<0.1%); by contrast, several proteins had *cis*-h^2^ approaching 100% (>99.9% for IL6R and PILRB), with 354 (12%) proteins having values >10%. A comparison of these *cis*-h^2^ estimates with those instead derived using association statistics for lead *cis* variants (using the formula 2pqβ^2^) showed generally good agreement (*r*=0.81) (**Supplementary** Figure 4C). By contrast, consistency between these methods was much poorer for *trans*-h^2^ (*r*=0.4, **Supplementary** Figure 4B), with genome-wide measures giving larger estimates; this presumably reflects contributions to estimated h^2^ at loci which failed to reach genome-wide significance. Comparing CKB and UKB h^2^ estimates for autosomal proteins, there were substantial differences between the two studies for many proteins; nevertheless, overall there were strong correlations for both total (*r*=0.64) and *cis*-h^2^ (*r*=0.74) (**Supplementary** Figure 5).

Many proteins had multiple pQTLs and/or conditionally independent associations (Figure 2D). Among the 1,784 proteins with at least one pQTL, the median number of conditionally independent associations was 2 (range 1 to 11), with 36% of proteins having a single and 15% having at least 5 associations. The proteins with the largest number of independent signals were CDHR5, CNTN5, CCL24, and MICA_MICB (11 each), with the majority for each being *cis* associations (35 *cis*, 9 *trans* overall) (Figure 2D). Conversely, as illustrated in Figure 2A, many loci are associated with levels of multiple proteins. Merging physically-overlapping loci across GWAS of all proteins, we identified 1,200 separate genomic regions associated with one or more proteins, including 424 regions containing only *cis*-pQTLs, 508 loci which were exclusively *trans*-pQTLs, and 268 which were both *cis*- and *trans-*pQTLs (**Supplementary Table 9**). There were 449 genomic regions associated with at least 2 different proteins, of which 45 were associated with ≥10 proteins (Figure 2E). The region showing the most extensive pleiotropy was chr9:132820126-137359331, a region of extended linkage disequilibrium (LD) which includes the ABO blood group locus and which was associated with 212 proteins. The next most pleiotropic regions were chr6:23826893-38728446, again featuring extended LD and spanning the HLA region (associated with 113 proteins), followed by chr19:50740785-55718384 (which includes the *KIR* gene cluster, associated with 60 proteins), chr19:48067663-49950310 (*NTN5* and *FUT2*, 51 proteins), and chr12:6373483-7813147 (*CD4* and *CD27*, 47 proteins). Pleiotropy was not restricted to *trans*- pQTLs, with 97 loci associated only in *cis* but with more than one protein (**Supplementary Table 9**). In particular, pQTLs with identical sentinel variants were associated in *cis* with 6 pairs of different but closely-related proteins (**Supplementary Table 4**) (e.g. AMY2A/AMY2B, C1R/C1S, NPPB/NT-proBNP).

### Fine-mapping

Inclusion of low frequency variants (>0.25%) in 2,923 parallel GWAS potentially results in an inflated Type I error^32,33^. To ensure downstream analyses used only high-confidence pQTLs, while avoiding highly-conservative Bonferroni correction as previously applied in UKB-PPP^21^, we applied an adjusted significance threshold of 2.497x10^-9^. This was derived by using a significance threshold of 5x10^-9^ suggested to be appropriate when including low frequency variants^32,33^, with further application of a hierarchical Benjamini-Hochberg procedure which effectively controls false-discovery for multiple GWAS applied to many phenotypes in parallel^34^. Comparison of effect sizes for lead variants in CKB with those reported for UKB-PPP shows that this adjustment leads to exclusion of several signals which pass the P<5x10^-8^ threshold in CKB but which have null associations in UKB-PPP (**Supplementary** Figure 6). At this revised threshold, 2,233 pQTLs were identified for 1,461 proteins, the large majority of excluded signals being *trans*-pQTLs with small effect sizes – only 79 *cis*-pQTLs were excluded, leaving a total of 1,233 *cis*-pQTLs for 1,201 proteins.

Fine-mapping at these 2,233 pQTLs identified 3,475 95% credible sets (CSs) (**Supplementary Tables 10 and 11**). For 21 pQTLs (1 *cis* and 20 *trans*), no CS was identified at a 95% posterior inclusion probability (PIP) threshold. CSs contained a median of 5 variants (mean 16, range 1 to 553), with 852 CSs (25%) fine-mapped to a single variant. Overall, *cis*-pQTLs had better resolution than *trans*-pQTLs (*cis*: median 4, mean 13 variants per CS; *trans*: median 10, mean 24), and more were mapped to a single variant CS (*cis*: N=658, 29%; *trans*: N=194, 17%). In a comparison of exact conditional analysis and fine-mapping at these 2,212 pQTLs, the two methods identified approximately the same number of association signals (3,674 vs 3,475 respectively) and 83% of lead variants from the conditional analyses were included within CSs. In subsequent analyses, for each CS we treated the variant with largest PIP as the lead variant; 81% of these were identical to variants identified in the corresponding exact conditional analysis.

### Characteristics of pQTL lead variants

For *cis*-pQTLs, 1266 (55%) of the 2,307 lead CS variants were within the transcript for the target (i.e. assayed) protein, although these were predominantly in non-coding regions (Figure 3A). Among those located in the coding region, >90% (407/448) were non-synonymous (i.e. protein altering variants [PAVs] affecting the amino-acid sequence of the translated protein, e.g. stop-gain, missense, insertion/deletion). These included 213 CSs consisting of a single missense variant, which might potentially be rare variants giving rise to measurement artefacts^35^, but only 23 of these were the sentinel variant (i.e. most strongly-associated) at that locus, and only 12 of these had a MAF<0.05 in either EAS or EUR. The remaining *cis*-pQTL lead variants were evenly distributed between intergenic (presumably regulatory) regions (541, 23%), and transcripts for genes other than the target gene (500, 22%), although these very rarely lay within coding regions. By contrast, the large majority of lead CS variants for *trans*-pQTLs lay in non-coding regions, with only 252/1,168 (22%) located in coding regions of gene transcripts, of which 195 were PAVs.

**Figure 3.**
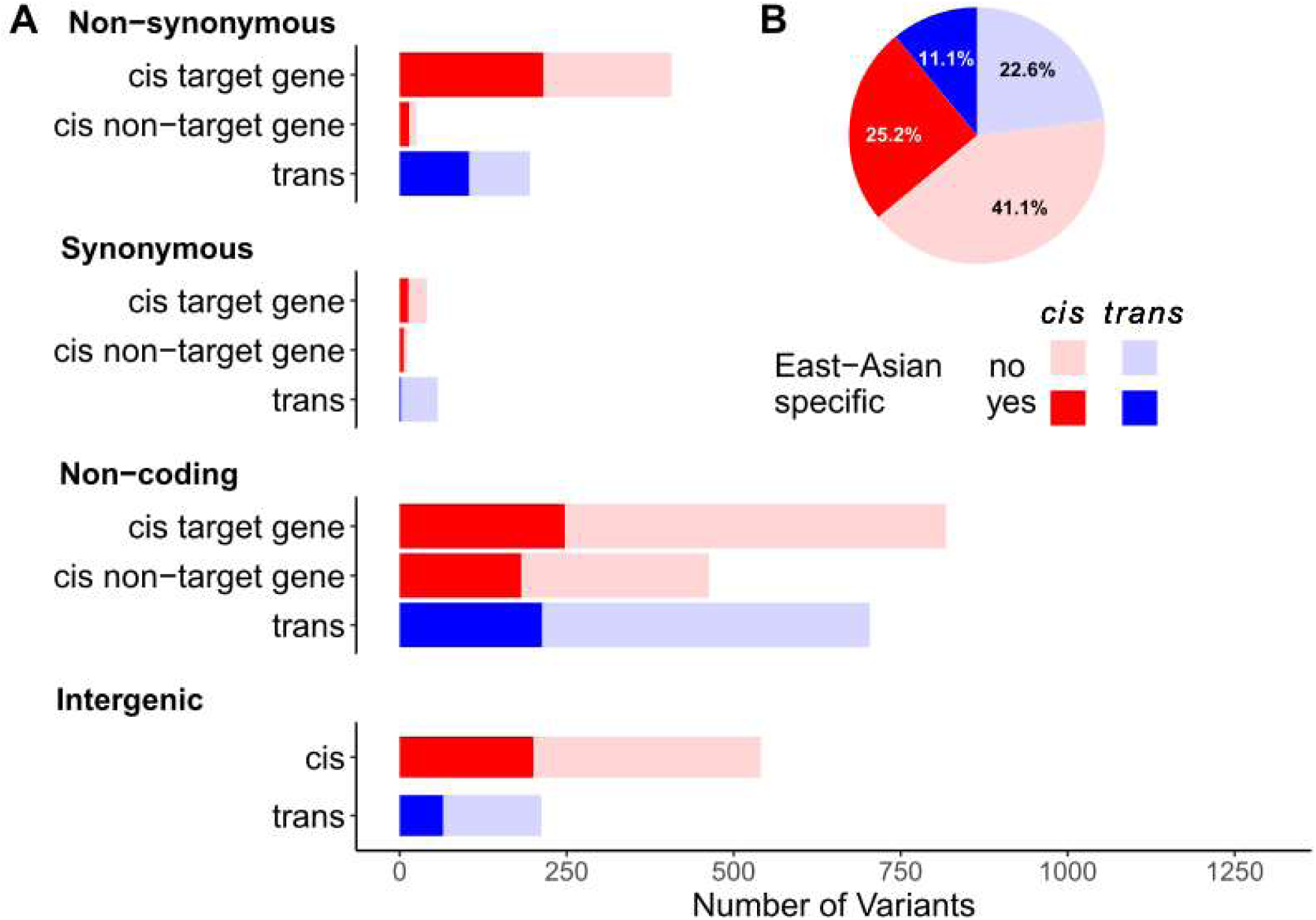
Location, impact, and novelty of pQTLs. (A) Functional annotation of lead variants from fine-mapped credible sets. Variants are categorised according to their whether they lie within a transcript encoding the assayed (target) or other protein, and their resulting impact on protein coding. **(B) EAS-specific pQTL associations**. Proportion of all fine-mapped credible sets for which there is evidence that they correspond to an association identified in UKB-PPP.

In addition to 610/3,475 (18%) pQTLs with lead variants being PAVs, a further 1,229 association signals included at least one PAV in their CS, of which 830 (68%) were *cis*-pQTLs. Thus, a total of 1,262 *cis*- and 577 *trans*-pQTL signals might potentially be due to PAVs (**Supplementary Table 11**). However, inspection of the proteins whose levels had multimodal distributions indicated that only a minority of these might be accounted for by measurement artefacts arising from benign changes to the protein. Only 12 such proteins had a *cis* CS including a PAV, and only 7 of these (ABO, CD300LF, FOLR3, IFNGR2, KLK12, PNLIPRP2, QPCT) had small CSs indicative of variants with large effect size (**Supplementary Table 11**).

### Comparison with European ancestry pQTLs

Of the 2,233 pQTLs identified in CKB at the adjusted P<2.497x10^-9^ significance threshold, there were 153, including 12 *cis*-pQTLs, for which no association was reported anywhere in that genomic region in the corresponding UKB-PPP discovery GWAS (**Supplementary Table 4**). For 7 proteins with at least one pQTL in CKB, including 3 (ALDH2, GLIPR1, SPACA5) with a *cis*-pQTL, there were none reported anywhere in the genome in UKB-PPP. Furthermore, only 317 (193 *cis*) sentinel variants were identical between CKB and the corresponding UKB-PPP discovery GWAS, with a further 570 being in very strong LD (r^2^>0.95; **Supplementary Table 4**). Despite this, where reported by UKB-PPP, 1,625/1,803 CKB sentinel variants were associated at genome-wide significance (P<5x10^-8^) (**Supplementary Table 12**). Conversely, 19% (430/2,233) of CKB sentinel variants were not included at all in the UKB-PPP study; in part this may be due to the exclusion of low frequency and poorly-imputed variants from the UKB-PPP analysis – 82% (354/430) have low allele frequency (MAF<0.01) in EUR (**Supplementary Table 4**).

Comparison of effect sizes in CKB and UKB-PPP showed overall consistency for both CKB and UKB-PPP sentinel variants **(Supplementary** Figure 6), but nevertheless there were appreciable differences between the 2 studies. For both sets of variants, effect sizes tended to be larger in the discovery cohort. perhaps reflecting winner’s curse, but even after accounting for this systematic difference there was appreciable heterogeneity of effect size between the two studies for a substantial proportion of pQTLs (**Supplementary Tables 12-13**). Of 1,803 CKB sentinel variants for which summary statistics were available in UKB-PPP, 1,049 (58%) had significantly different effect sizes at 5% FDR. By contrast, 15% (1,630/10,824) of UKB-PPP sentinel variants showed heterogeneity of effect size compared with CKB, but this likely reflects limited power in CKB to detect small-effect *trans*-pQTLs – if only *cis*-pQTLs were considered, there was heterogeneity for 44% (607/1,392) of UKB-PPP variants.

To more systematically assess the extent to which pQTL associations in CKB were distinct from those reported in UKB-PPP, we further tested the LD between the lead variants in each CS in CKB with those for the reported CSs for the same protein in UKB-PPP. Overall, even at a permissive LD threshold of r^2^≥0.8 (assessed in both EAS and EUR LD reference panels), nearly half of all lead CSs identified in CKB (1,491/3,475, 43%) were not in LD with any variant in CSs from UKB-PPP (**Supplementary Table 11**), with *cis* and *trans* signals equally likely to be identified as potentially novel. This substantial proportion of apparently novel associations was only partly accounted for by CKB lead variants being absent from the UKB-PPP analysis and instead imperfectly tagged by more common variants – 101 (7%) lead variants for CKB-specific signals have low MAF (0.001-0.01) in EUR populations but are potentially responsible for an observed association signal in UKB at a more common variant (UKB D’>0.95). Conversely, 629 (41%) such lead variants are extremely rare in EUR populations (MAF<0.001) and thus represent high-confidence EAS-specific signals. Consistent with this, the lead variants of CSs not in LD with lead UKB-PPP variants had appreciably lower MAF than the remainder in both CKB (mean MAF 0.17 and 0.25, respectively) and non-Finnish Europeans (0.11 and 0.23) (**Supplementary Table 11, Supplementary** Figure 7). Likewise, if restricting to loci identified at the more stringent Bonferroni-adjusted significance threshold used in UKB-PPP, similar proportions of *cis* and *trans* associations in CKB (42% and 38%, respectively) appeared distinct from those in UKB-PPP. Of particular note, the *cis*-pQTLs for 256 proteins had no CSs that overlapped with those identified in UKB-PPP; only 38 of these proteins had a *cis*-pQTL whose effect size was not significantly different from that reported by UKB-PPP (**Supplementary Table 12**).

As an alternative appoach to assessing potential novelty of associations, we assessed whether CSs identified in CKB had any variant in common with any CS reported for that protein by UKB-PPP. Overall, by this criterion 48% (1,677/3,475) of CKB associations were novel compared to UKB-PPP pQTLs (1,201 *cis*, 52%; 476 *trans*, 41%) (**Supplementary Table 11, Supplementary** Figure 8). There was good agreement with the LD-based approach, with 1,366 (82%) of these signals also being identified as not being in LD with UKB-PPP lead variants (**Supplementary Table 11, Supplementary** Figure 9). Again, only a small proportion (107, 6%) of this inferred signal novelty can be accounted for by lead variants which are low-frequency in EUR but potentially tagged by more common variants identified as associated in the UKB-PPP analysis. When restricting to loci passing a Bonferroni-adjusted significance threshold, the proportion of “novel” association signals was again similar (52% and 37% for *cis* and *trans*, respectively). By this approach, the *cis*-pQTLs for 359 proteins had no associations in common with UKB-PPP, including 225/256 of those identified by the LD approach.

We also used cross-ancestry fine-mapping with SuSIEx^36^ to identify associations present in both CKB (PIP>0.8) and UKB-PPP (PIP>0.8) (**Supplementary Table 14**). Cross-ancestry credible sets were identified in only 904/2,233 pQTLs, with only 1,015/3,475 CKB lead variants (29%) included in the CSs for such associations. There was strong agreement with the methods used above, with 89% and 85% of SuSIEx-identified cross-ancestry associations also being identified according to LD or CS overlap, respectively. Consequently, 36% (1,261/3,475 of CKB pQTL associations (877 *cis* and 384 *trans*) were identified as distinct from those in UKB-PPP according to all 3 methods, the locations and characteristics of lead variants for these associations having a similar pattern to the remainder (Figure 3). There were 205 proteins with *cis-*pQTLs for which there was no CS showed evidence of being shared across ancestries.

### Phenome-wide scan and Mendelian randomisation analyses

We searched GWAS Catalog^37^ for associations with sentinel variants for the 1,228 unique *cis*-pQTLs passing the 2.497x10^-9^ threshold (including multiple sentinel variants for proteins with >1 non-overlapping *cis*-pQTLs, and 5 variants associated with 2 different proteins), using the best available proxy (LD *r*^2^ > 0.8) where necessary. At the default search threshold of P<5x10^-6^, a total of 377 pQTL- trait and 103 pQTL-disease (Figure 4**, Supplementary Table 14**) associations were identified in studies using exclusively EAS populations. Out of the 480 identified associations, 417 surpassed a Bonferroni-adjusted threshold of P<1.595x10^-8^ (0.05 / [1,228 pQTLs x 2,552 traits]), comprising 81 protein-disease pairs and 336 protein-phenotype associations. Notably, significant disease associations with 12 out of 47 proteins were based on distinct *cis*-pQTLs from those previously identified in UKB-PPP – i.e. none of the lead variants at that locus were in LD with those reported by UKB-PPP – and for a further 15 proteins at least one *cis*-pQTL lead variant was not in LD with those reported in UKB-PPP. We also searched 3,215 traits with multi-ancestry GWAS that included EAS populations and identified 946 protein-phenotype and 108 protein-disease pairs (**Supplementary Table 14**) with P<1.26x10^-8^. However, the multi-ancestry nature of such studies complicates interpretation of these results, and these were not explored further.

**Figure 4.**
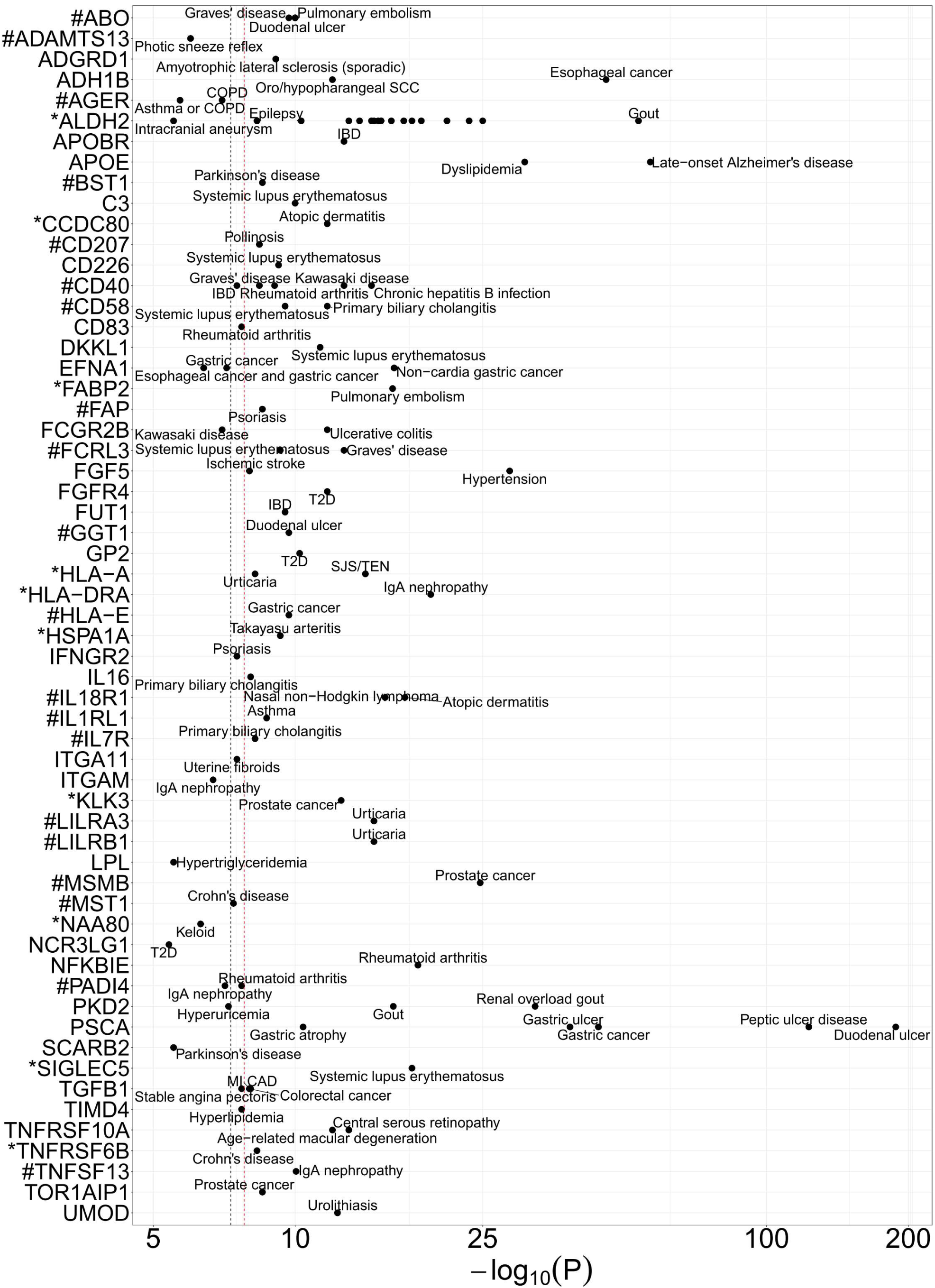
**pQTL-disease associations in East Asian studies from GWAS Catalog**. Associations are shown for sentinel *cis*-pQTLs or their strongest available proxies (LD r^2^>0.8). Lines denote genome-wide (black, P=5x10^-8^) and Bonferroni-adjusted (red, P=1.59x10^-8^) levels of significance. Asterisks and hashtags denote *cis*-pQTLs whose constituent *cis* credible sets were all or partly identified as East Asian-specific, respectively.

To assess the strength of the associated proteins’ putative effects on disease risk, we next performed two-sample MR analyses for 59 of the most robustly-associated protein-disease pairs. The potential for confounding by LD was reduced by limiting to disease-associated variants which were the same as or in very strong LD (r^2^>0.95) with the corresponding pQTL sentinel variant, and we retained only the most strongly-associated where there were multiple similar disease associations (e.g. ALDH2 with coronary heart disease, myocardial infarction, and unstable angina) (Figure 5, **Supplementary Table 15**). Using effect size estimates and effect alleles from the relevant EAS-specific studies, many proteins had a large apparent effect on the associated diseases, with a genetically-predicted 1 SD change in protein level being associated with at least a 2-fold increase or 0.5-fold decrease in risk for 19 protein-disease pairs. Particularly strong effects on disease risk were observed for ADH1B on oesophageal cancer (OR=33.1 [95%CI 20.5-53.6] per 1 SD higher protein level) and oro/hypopharyngeal squamous cell carcinoma (19.4 [8.5-44.2]), and for ALDH2 on multiple outcomes including oesophageal cancer (0.27 [0.21-0.35]), gout (6.0 [4.8-7.5]), and epilepsy (2.7 [1.9-3.8]). For these two proteins, the apparent effect sizes may be artefactually high since in both cases the lead variant is associated not only with altered protein levels but also with changes in protein function. Moreover, their associations with disease risk are believed to be mediated mostly through their effects relating to alcohol consumption^38^, rather than having direct causal effects.

**Figure 5.**
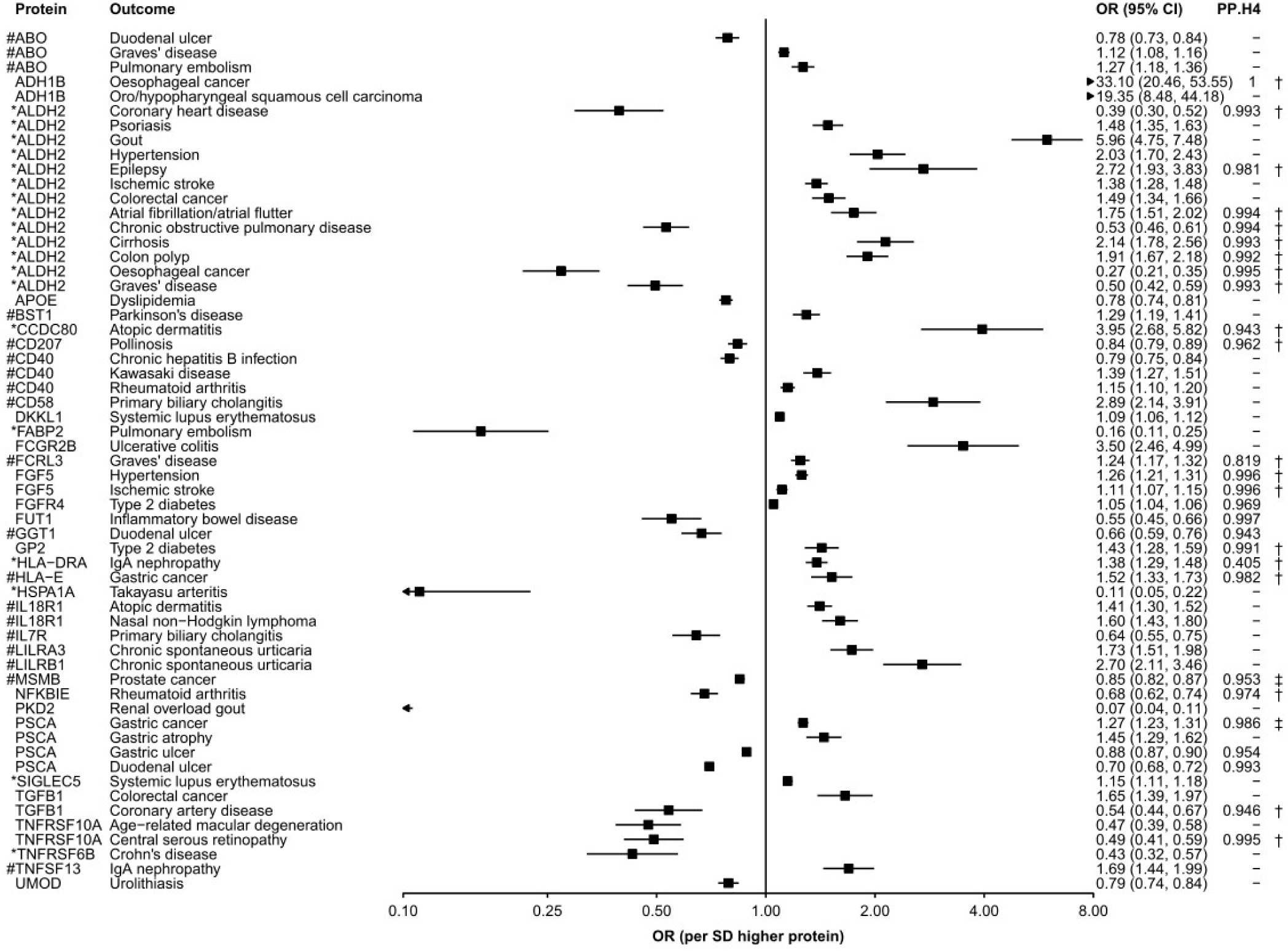
Estimation of protein effects on disease risk. Wald ratio effect estimates are shown for 1SD higher protein levels. Asterisks and hashtags denote *cis*-pQTLs whose constituent *cis* credible sets were all or partly identified as East Asian-specific, respectively. The posterior probability that pQTL and disease associations colocalise using (†) coloc.abf or (‡) coloc.susie are given for all analyses for which PP.H3+PP.H4>0.5.

Many other proteins, however, may have direct effects on disease risk, suggesting that they represent potential drug targets for prevention or treatment. These include multiple proteins for which the *cis*- pQTL associations were entirely distinct from any identified in UKB-PPP: for instance, higher levels of CCDC80, CD58, and LILRB1 were associated with substantially increased risk of atopic dermatitis (3.95 [2.68-5.82]), primary biliary cholangitis (2.9 [2.1-3.9]) and chronic spontaneous urticaria (2.7 [2.1- 3.5]), respectively, while FUT1, HSPA1A, and TNFRSF6B were associated with decreased risk of inflammatory bowel disease (0.55 [0.45-0.66]), Takayasu arteritis (0.11 [0.05-0.22]), and Crohn’s disease (0.43 [0.32-0.57]), respectively. For several of the protein-disease associations, the genomic region containing the associated *cis*-pQTL also included *cis*-pQTLs for one or more other proteins (**Supplementary Table 9**). In all cases, however, none of the other proteins in the region showed any association with the corresponding disease, with the exception of LILRA3 and LILRB1, which have identical *cis*-pQTLs and were both associated with chronic spontaneous urticaria.

We compared the results of this analysis with a similar search of GWAS Catalog performed using UKB-PPP *cis*-pQTLs^39^. For more than half of these protein-disease associations, no equivalent or similar association was identified in the previous study; even excluding the 13 associations with ALDH2 (which has a well-established EAS-specific functional variant), approximately half (21 out of 46) of the protein-disease pairs were not previously identified based on pQTLs and disease GWAS from EUR populations (**Table 1**). Of the 7 proteins for which CKB *cis*-pQTLs were inferred to be entirely distinct from those identified in UKB-PPP by three separate methods (LD, CS overlap, SuSIEx), 6 had one or more disease associations not identified in the EUR study. In addition, a further 7 proteins with at least one putative EAS-specific *cis* association had disease associations not previously reported.

**Table 1.**
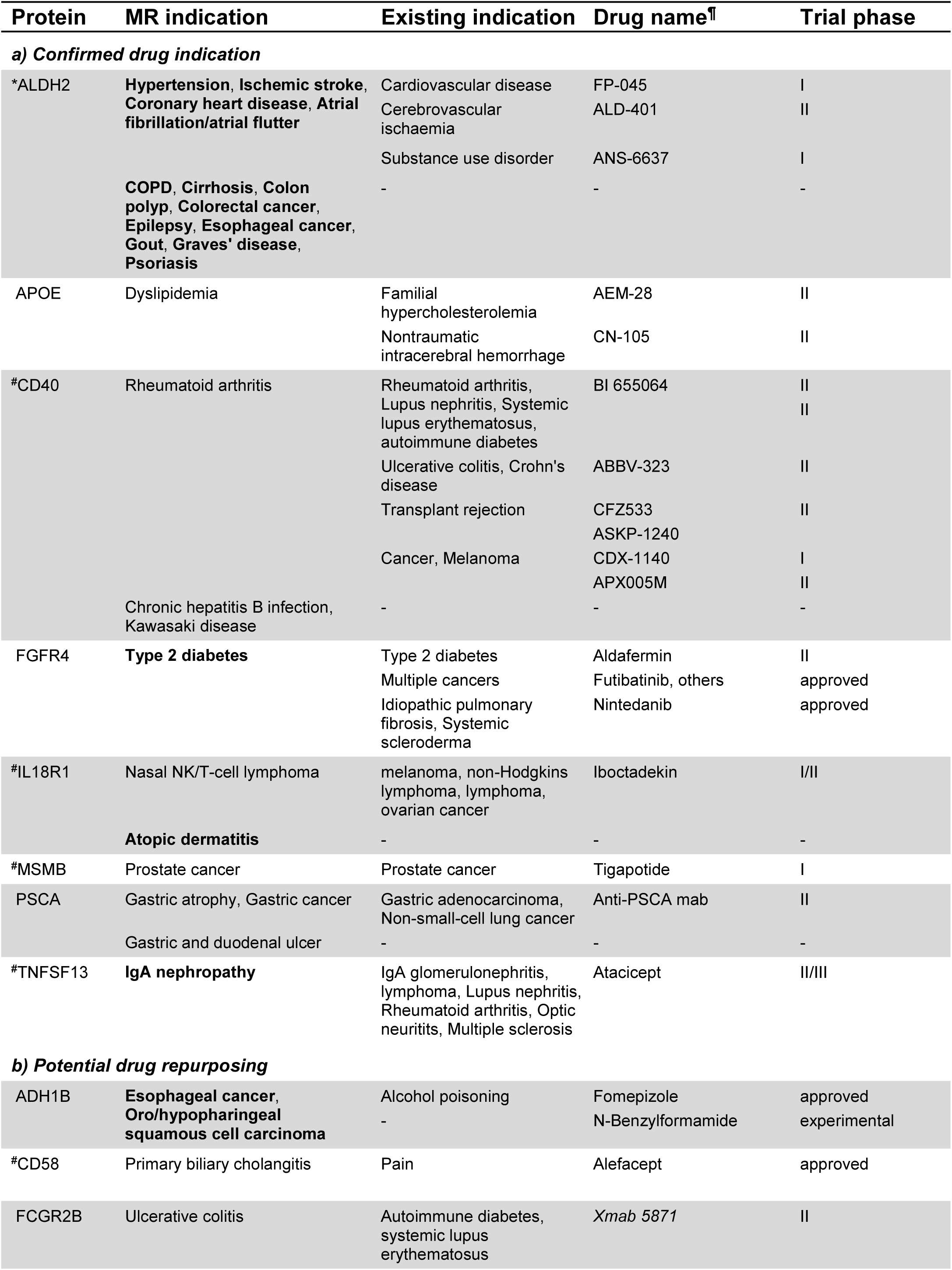
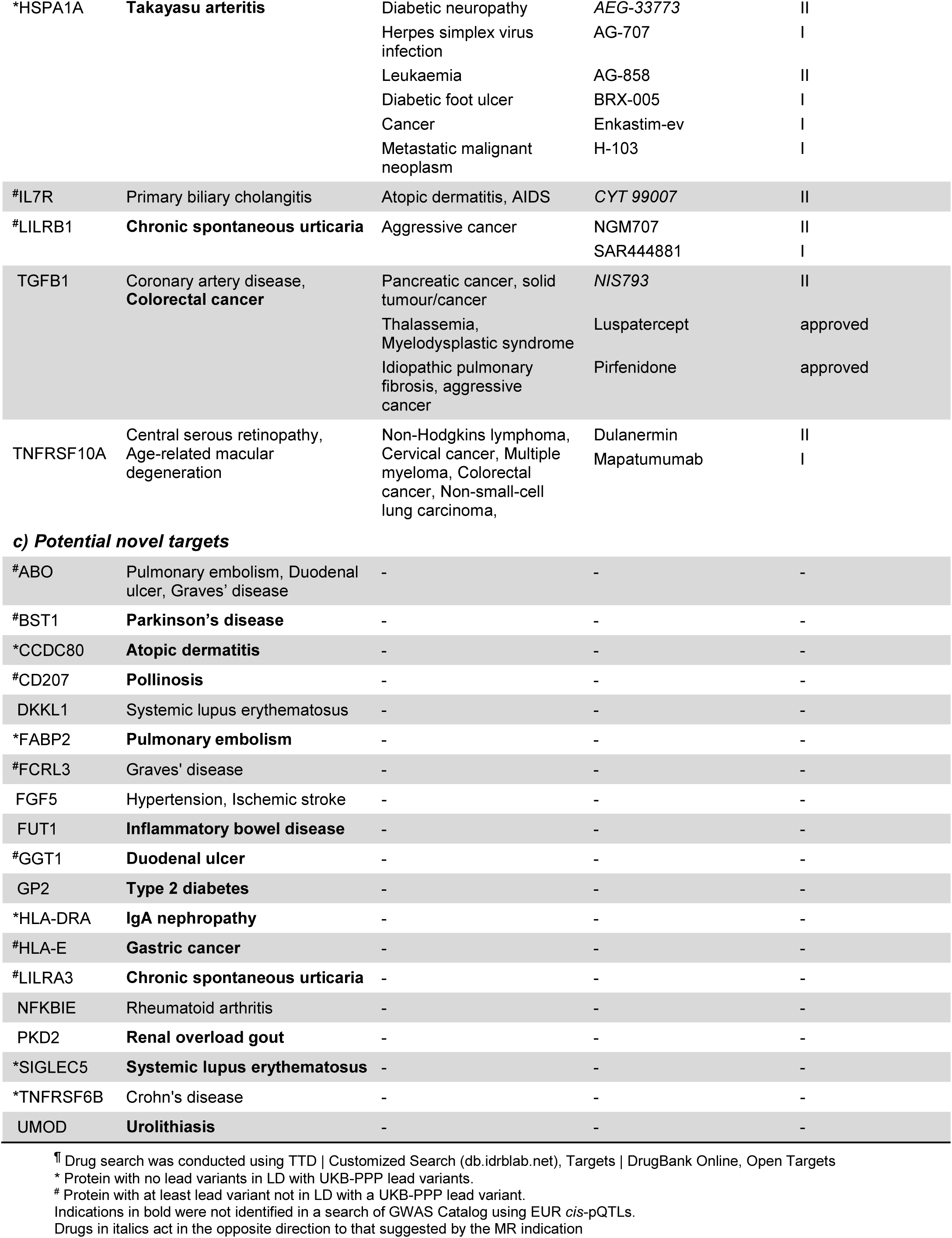
Drugs and main indications relevant to disease-associated proteins.

We further assessed the causal relationship of 36 of the 59 protein-disease pairs by testing colocalisation of the pQTL association signal with summary statistics for the corresponding disease, where available either at GWAS Catalog or from Japanese^40^ or Korean^41^ biobanks (Figure 5, **Supplementary Table 16**). There was strong evidence for colocalisation (PP.H4>0.9) for 26 out of 28 disease-protein pairs, although for MSMB-prostate cancer and PSCA-gastric cancer this was only observed when using SuSIE-coloc to test colocalisation of separate association signals, perhaps reflecting differences in protein expression in disease-relevant tissues compared to blood; the results were inconclusive (PP.H3+PP.H4 <0.5) for the remaining 8 protein-disease pairs. In no instance was there strong evidence to refute a causal relationship between protein level and disease risk. For the remaining protein-disease associations, we were unable to access disease summary statistics which were sufficiently well-powered for colocalisation analysis.

### Potential drug target and repurposing opportunities

We searched the Therapeutic Target Database^42^, DrugBank^43^, and Open Targets^44^ to identify existing drugs targeting the 35 separate proteins from our MR analysis, and their corresponding disease indications (**Table 1**). Of these, 16 are targeted by at least one drug either approved, under investigation in clinical trials, or reported in the literature, and in 8 cases a drug targeting the protein was a potential treatment for the disease identified in our analyses. For 4 of these proteins (ALDH2, CD40, IL18R1, PSCA), we identified additional disease associations distinct from the current intended indications, and for a further 8 proteins the drugs which target them are again for indications different from the associated disease(s). Notably, we did not identify any drugs targeting 19 proteins, including 5 of those for which CKB *cis*-pQTLs were distinct from those in UKB-PPP and 13 protein-disease associations not identified in the equivalent UKB-PPP analysis.

## Discussion

There have been only limited genetic analyses of the human proteome in EAS populations^15,29,30^. In this study, high-throughput Olink Explore 3072 PEA proteomic assays in the largest EAS sample to date have enabled detailed characterisation of the genetic architecture of 2,923 plasma proteins in Chinese adults, and comparison with that in a European population. We identified 4,659 independent pQTL association signals for 1,784 proteins at genome-wide significance, including 2,577 independent associations within 1Mbp of the structural gene for the corresponding protein. We replicated the large majority of pQTLs previously identified in EAS populations, and identified a large number of additional pQTLs, especially *trans* associations. In addition, the increased power from our larger sample size enabled greatly improved fine-mapping; CSs with PIP>0.95 were identified for 97% of associations, including 25% of signals resolved to a single variant.

In the recently-reported UKB-PPP GWAS of >50K UK Biobank participants (predominately EUR) using the same Olink panel^21^, pQTLs were identified for 83% of the proteins, with at least one *cis*-pQTL for 70% of the proteins^21^. Notably, despite having an approximately ∼10-fold smaller discovery sample size, we identified pQTLs at genome-wide significance for 61% of the proteins. Even at a more stringent adjusted threshold (P<2.497x10^-9^), we identified 2,233 pQTLs for 1,461 proteins (50%), including 153 CKB pQTLs for 140 proteins at genomic locations where no corresponding association was reported by UKB-PPP. Furthermore, a systematic comparison of CSs from the two studies showed that the lead variants for 43% of CKB CSs were not in appreciable LD (*r*^2^ > 0.8) with the corresponding associations in UKB-PPP. Similarly, 48% of CKB CSs had no overlap with CSs reported by UKB-PPP, and only 29% of CKB pQTLs colocalised across the two studies, such that 36% of CKB pQTL associations appeared distinct from those in UKB-PPP according to all 3 methods. Thus, it is clear that there are substantial ancestry differences in the genetic architecture of the proteome.

Despite these differences, the two studies identify similar large-scale features of the genetic architecture of the proteome; for instance, there is substantial pleiotropy for loci at ABO, KIR, FUT2, and within the HLA region. Moreover, in line with the findings in UKB-PPP^21^, there were substantial differences in the patterns of effect sizes for *cis*- and *trans*-pQTL minor alleles, with the minor alleles of *cis*-pQTL lead variants being more likely to be associated with larger effect sizes and with lower measured protein levels. It seems likely that this relates to the high proportion of both *cis*-pQTL CSs (1,262/2,307, 55%) and *trans*-pQTL CSs (577/1,168, 49%) that include at least one PAV. Variants leading to protein truncation or structural changes are likely to reduce rather than increase levels of that protein; thus, there will be a consistent direction of effect for such *cis*-pQTLs, by contrast with the regulatory effects of *trans* loci which may either increase or decrease levels of the target protein.

The benefits of multi-ancestry proteogenomic studies are demonstrated from the results of our phenome-wide screen of association signals in GWAS Catalog^37^. Using GWAS specific to EAS populations, which are far fewer and usually less well-powered than those available for EUR, we identified 417 distinct significant associations of *cis*-pQTLs with phenotypes and diseases. These included 19 disease-associated proteins not targeted by any drugs currently in development, and a further 12 proteins targeted by drugs in development but with at least one associated disease which is not an indication for those drugs. Of these, 6 proteins had *cis*-pQTLs which were distinct from those identified in UKB-PPP. Furthermore, a comparison with results from a similar analysis using data from EUR populations found that disease associations for 21 proteins were not identified using UKB-PPP *cis*-pQTLs, despite the larger number of available *cis*-pQTLs and GWAS (4,439 disease/phenotype associations identified for a total of 646 proteins)^39^.

Use of *cis-*pQTLs, as genetic instruments in two-sample MR^45,46^, is becoming established as a powerful approach for the identification of possible causal effects of protein levels on diseases and phenotypes, which can inform drug development. The identification of *cis*-pQTLs on the basis of their proximity to the relevant structural gene means that their primary mode of action is likely to be a direct effect on protein levels, especially in the case of PAVs. Thus, association of such variants with additional phenotypes or diseases are considered likely to reflect a causal relationship, suggesting a mechanistic basis for previously-unexplained GWAS signals for disease.

Although potentially vulnerable to genetic confounding due to LD^19^, epitope effects (in which protein sequence changes due to a PAV disrupt the Olink assay)^35^ or co-regulation with other nearby genes, we found no evidence for these being issues for *cis*-pQTLs in our analysis: in 27/28 cases where sufficiently well-powered summary statistics were available, colocalisation between CKB protein GWAS and disease GWAS supported a causal link between protein levels and disease risk; despite most of our disease associations being in genomic regions harbouring multiple *cis*-pQTLs, with one exception (LILRA3 and LILRB1) where no pQTLs for other nearby proteins were associated with the same disease; and where observed the co-regulation in CKB was for pairs of very closely-related proteins with similar functions (LILRA3/LILRB1 being the only pair of disease associated proteins). By contrast, *trans*-pQTLs are by their nature vulnerable to misleading findings due to pleiotropy, since their effects on protein levels are likely to also reflect modulation of other (often un-assayed) proteins, mediated by proteins for which they are also *cis*-pQTLs. In our study, 66% of *trans*-pQTLs were associated with more than one assayed protein, hence, we did not use *trans*-pQTLs in our primary analyses.

The utility of *cis*-pQTLs for the identification of potential drug targets is illustrated by the directionally-consistent associations between several existing targets and one or more of the indications for which drugs are currently in development: there are inferred protective effects of APOE and MSMB on dyslipidemia and prostate cancer, respectively, and synthetic peptides mimicking the action of these proteins are intended for use in treatment of these diseases^47,48^; conversely, association of CD40, PSCA, and TNFSF13 with increased risk of various diseases is reflected in drugs intended to inactivate or down-regulate these proteins. In addition, we identified associations of CD40 and PSCA with additional diseases not currently intended for treatment by the drugs in development. However, although these might represent repurposing opportunities, their respective associations with chronic hepatitis B infection and gastric/duodenal ulcers are in the opposite directions to those for other diseases, so that existing drugs targeting these proteins may not be of benefit for these indications.

The majority of the above associations were also identified in the recent similar analysis using UKB-PPP pQTLs^39^, perhaps reflecting the strong bias towards EUR populations in the evidence base underpinning drug development. By contrast, less than half of the potential repurposing opportunities and novel indications were identified in the previous analyses. Most of the novel associations were for CKB *cis*-pQTLs distinct from those identified in UKB-PPP, emphasising the value of ancestry diversity. In other cases, the same *cis*-pQTL was identified in both ancestries yet disease associations were only identified in analyses based on CKB pQTLs and disease GWAS from EAS populations, In some cases this may reflect much smaller MAF in EUR populations, presumably limiting power to detect disease associations, for example the association of the *cis*-pQTL for ADH1B with oesophageal cancer (MAF=0.32 and 0.04 in EAS and EUR, respectively). In other cases it may reflect diseases or associations observed or analysed primarily in EAS populations: Takayasu arteritis commonly presents in East and South Asian women before the age of 40^49^ so that the identification of this possible drug target indication would have been unlikely in the absence of ancestry-diverse studies; an association with type 2 diabetes at the GP2 locus has only been observed in EAS and not EUR populations^50,51^, perhaps because the sentinel variant is 4-fold more common in EAS; and while the role of UMOD in kidney function and kidney disease is well-established, the only studies of urolithiasis in EUR have focussed on protein altering or rare copy number variants^52,53^,

These potential novel drug target opportunities were identified based on proteomics measures in only 4,000 Chinese adults, and there would be substantial benefits from increasing sample size, which would, as evidenced in UKB-PPP, lead to the identification of many additional pQTLs, particularly *trans*- pQTLs and corresponding associations with disease^21^. In addition, more pQTLs will be identified as the coverage of measured proteins increases, not only on the Olink platform but also using other technologies. Additional *cis*-pQTLs and disease associations are identified using the SomaScan platform, which includes many proteins not included on the Olink EXPLORE panel^39,54^, while mass- spectrometry proteomics of 304 proteins in 2,958 Han Chinese, with minimal overlap with the Olink EXPLORE panel, also identified many potential protein-disease associations^30^.

One important limitation of this and similar proteomics studies is that plasma protein levels do not necessarily reflect levels of the protein in the disease-relevant tissue, and are frequently not normally-distributed, so that protein effects on disease may be mis-estimated. Furthermore, functional alterations to a protein (e.g. arising from missense mutations) which do not affect the abundance or conformation of a protein might remain undetected, potentially masking associations between protein and diseases. Conversely, protein altering variants, which are potentially responsible for up to 55% of *cis*-pQTL associations in this study, can have epitope effects which alter the results of the protein assay without influencing protein levels or function. While the consistent support for protein-disease associations by colocalisation suggests that these issues are rare, the novel findings from this study will require further confirmation. In this respect, another limitation of this study is the lack of comprehensive EAS-specific resources (e.g. multi-tissue eQTL and sQTL data) to strengthen the evidence for potential drug target opportunities.

In conclusion, the present study is the largest proteome GWAS to date in an EAS population, in terms of both the number of proteins assayed and sample size. Our analyses highlighted distinct ancestral differences in the genetic architecture of plasma proteomics, with approaching 50% of pQTLs being distinct from those previously identified in EUR populations. The identification of many novel protein-disease associations demonstrates that extending genome-wide plasma proteomic analyses to non-European ancestry populations has great potential for the identification of novel drug targets for major diseases and the associated traits.

## Acknowledgments

The chief acknowledgment is to the study participants, the members of the survey teams in each of the 10 regional centres, and the project development and management teams based at Beijing, Oxford and the 10 regional centres. We thank Sarah Clark, Martin Radley, and Mike Hill at CTSU, Oxford, for assisting with the planning and organisation of the proteomics measurements. Access to UK Biobank data was under project 50474.

## Author contributions

RGW & ZC designed the study, AP, SS & RGW analysed the data and drafted the manuscript. SM, AE & AI further analysed or annotated the data. RGW, IM, RC and ZM supervised the study. KL, CK, NW, DA, HF, YC, HD, DVS, PP, JL, CY, DS, JC, LL, and DB collected and/or prepared the data. All authors including RP, RC and MH reviewed the manuscript.

## Ethics

Ethical approval was obtained from the Ethical Review Committee of the Chinese Centre for Disease Control and Prevention (Beijing, China, 005/2004) and the Oxford Tropical Research Ethics Committee, University of Oxford (UK, 025-04), and all participants provided written informed consent.

## Data availability

CKB non-genetic data are available and updated periodically for access by bona fide researchers. Details of the CKB Data Sharing Policy, data release schedules and data request application procedures are available at www.ckbiobank.org. Queries about data access should be addressed to. Proteomic data analysed in this study are available at https://doi.org/10.6084/m9.figshare.27931350. Access to individual participant genetic data is currently constrained by China’s Administrative Regulations on Human Genetic Resources, for which collaboration with CKB researchers is generally required, and which may be subject to separate regulatory approvals in China if it involves substantial sharing of unpublished data. Full proteogenomics summary statistics are available at GWAS Catalog [<u>accession codes pending</u>] and on the CKB PheWeb (pheweb.ckbiobank.org).

## Code availability

No custom code or algorithms were generated.

## License

This research was funded in whole, or in part, by the Wellcome Trust [212946/Z/18/Z, 202922/Z/16/Z, 104085/Z/14/Z, 088158/Z/09/Z]. For the purpose of Open Access, the author has applied a CC-BY public copyright licence to any Author Accepted Manuscript version arising from this submission.

## Methods

### Study population and design

The CKB study design and population have been previously reported^55–57^. Briefly, 512,891 Chinese adults aged 30-79 years were recruited from the general population in 5 urban and 5 rural areas in 2004-2008. Questionnaire data, physical measurements and blood samples were collected by trained health workers in local assessment centres. The characteristics of study participants included in this study are reported in Supplementary Table 1.

The present study design was a case-subcohort within the prospective CKB study. This included 1,951 incident cases of IHD (ICD-10: I21, I20-I25]) accrued during a 12-year follow-up prior to 1^st^ January 2019, and 2,026 sub-cohort participants. The IHD cases comprised incident IHD cases that had GWAS data and no prior history of CVD (i.e. IHD, stroke, transient ischaemic attack or rheumatic heart disease) and no use of statins at enrolment. The sub-cohort was randomly-selected from a population-representative subset of 69,353 participants with GWAS data^57^ and with no reported history of CVD at baseline. All participants in this study were unrelated, no pair of individuals having a KING-robust kinship estimator >0.05 as assessed using PLINK v1.9^58^.

### Proteomic assays

Proteomics measurements were as previously described^59^. Relative levels of 2,923 unique plasma proteins were measured in 3,977 participants using the Olink Explore 3072 PEA panel (Olink Inc, Uppsala, Sweden)^20^, expressed as NPX units on a log2 scale. Protein values below the lower limit of detection were included in analyses while those with QC or assay warnings were excluded. Data for 3 OLINK batch 1 outlier samples and 9 OLINK batch 2 outlier samples were excluded based on PCA of the protein levels (|Z-score|>5 for any of the first 10 PCs), giving sample sizes of 3,974 and 3,968 participants respectively. Protein measures were analysed as rank inverse normal transformed residuals following linear regression on sex, age, age^2^, and recruitment region (10-level categorical variable). Proteins with multi-modal distribution were identified by visual assessment of histograms of protein level distributions.

### Genomic data

Genotyping of CKB participants was as previously described^57^, with imputation as follows: genotypes of 531,565 variants passing QC in all 100,706 genotyped samples were converted to genome build 38 using CrossMap v0.6.1^60^, and were checked for consistency by reversing the process (“liftUnder”). Variants not mapped, mapped to different chromosomes, or not mapped back to the same locations after liftUnder, were excluded. The remaining 531,542 variants were prephased using SHAPEIT v4.2 (SHAPEIT v2.904 for chromosome X)^61^, and genotypes were imputed using (i) the TOPMed imputation server (release 2 v1.0.0)^62^ and (ii) the Westlake Biobank for Chinese imputation server^63^; the two sets of imputed data were merged, for each variant retaining the imputed genotypes with the higher imputation INFO score. Variants with INFO<0.3 or MAF=0 were excluded, giving a final imputed dataset of 17,933,159 variants with MAF>0.001. CKB allele frequencies were derived as the mean imputed dosage, and in non-Finnish Europeans were from gnomAD v3.1.2^64^.

### Genome-wide association analyses

Genome-wide association analyses were performed for the rank inverse normal transformed protein levels using BOLT-LMM v.2.3.4^65^ with 11 PCs and genotyping array version as covariates. Post GWAS filtering removed SNPs with effective minor allele count <20 (MAF * INFO score * 2 * N <20), corresponding to a minimum MAF of 0.0025.

pQTL loci were identified by LD clumping around association signals reaching genome-wide significance (P-value <5 x 10^-8^), using PLINK v1.9^58^ to identify variants in linkage disequilibrium (LD r^2^>0.05, P-value >0.05) within 5Mbp of the lead variant (or within 20Mbp for association signals near the HLA region, chr6:21744977-39074734^57^), based on LD from 72K unrelated CKB participants with imputed genotype probabilities converted to best-guess genotypes. Locus boundaries identified by clumping were extended by 10Kbp, and physically overlapping loci were merged. The sentinel associated variant for the locus was that with the lowest P-value with ties decided according to the largest absolute Z-score. Conditionally independent association signals within each merged locus were identified using exact step-wise conditional analysis by including the allele dosage of associated variants as covariates; conditional effect sizes at a locus were obtained by fitting a joint linear model in R that included all independently-associated variants. pQTLs were categorised as *cis* or *trans* according to whether the sentinel variant lay within 1Mbp of the structural gene for the assayed protein (**Supplementary Table 3**).

To determine whether inclusion of incident IHD cases may have biased the GWAS results, association of sentinel variants was assessed by multiple approaches: (i) Logistic regression in the full genotyped CKB dataset to assess if case status was associated with the variant genotype, adjusting for covariates (age, age^2^, sex, region, 11 PCs); (ii) tests using the R anova() function for improved model fit by addition of case ascertainment to a linear regression model for protein measurement associated with variant genotype and covariates age, age^2^, sex, region, and 11 PCs; and (iii) comparison of effect size (betas) and P-values from the same two linear regression models. Finally, we performed separate GWAS for each protein in the IHD cases and subcohort, compared the resulting sentinel variant effect sizes with those from our discovery GWAS (**Supplementary** Figure 10), and compared the discovery P-values with those resulting from meta-analysis of the IHD and subcohort strata (**Supplementary** Figure 11). In no case was there any indication of potential bias from inclusion of IHD cases.

### Significance threshold

To enable direct comparison of with the previous Japanese study^29^ and with UKB-PPP, initial identification of pQTLs and conditionally independent associations used a genome-wide significance threshold of *P*<5x10^-8^, with further application of a Bonferroni-adjusted threshold of P<1.71x10^-11^ (5x10^-8^/2,923 proteins). For the main analyses, adjustment for multiple testing across 2,923 GWAS followed a hierachical Benjamini-Hochberg procedure^34^, treating each GWAS as a “family” of tests. The Simes’s P-value^66^ was derived for each GWAS across all identified pQTLs, and the significance threshold was determined as that giving a Benjamini-Hochberg false discovery rate of 5x10^-9^, applied to the set of 2,923 Simes’s P-values (i.e. 2.497x10^-9^ = 5x10^-8^ x [1,460/2,923 GWAS with a Simes’s P <2.497x10^-9^]).

### Replication

Replication of CKB sentinel variants and of pQTLs from the previous Japanese study^29^ used available summary statistics (*cis*-pQTLs only). Where data were not available for sentinel variants, proxies were identified in the discovery study (r^2^ > 0.8 and P < 5 x 10^-6^). For comparison of effect sizes between CKB and UKB-PPP^21^, only sentinel variants were assessed to avoid complications arising from differing LD between EAS and EUR populations. Systematic differences in effect between the CKB and UKB-PPP studies were determined using the deming R package and heterogeneity according to Cochran’s Q was assessed after scaling to give equal effects.

### Heritability

Total narrow-sense autosomal heritability for each protein was estimated using *GCTA*^67^ using transformed protein level measures (i.e. including adjustment for the same covariates as in the GWAS analysis): *trans*-heritability was estimated by excluding SNPs in protein encoding regions (±1Mb) from the genetic relationship matrix (GRM); *cis*-heritability was estimated as the difference between total and *trans* heritabilities. The standard error for *cis* heritability was estimated as:

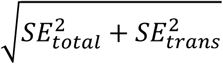

For proteins encoded on chrX, *cis h*^2^ was determined from conditionally independent association signals according to the formula:

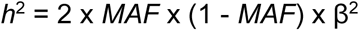

where β is the conditional effect size in SD units, with total *h*^2^ derived by adding to total autosomal *h*^2^. Heritability for all proteins was also determined in the same manner, as the sum across all single lead SNPs at each associated locus, with *cis*- and *trans-*heritability partitioned according to the corresponding pQTLs.

### Fine-mapping

Fine-mapping within clumped and merged pQTLs used SuSiE version 0.12.35^68^ to obtain CSs of likely causal variants with a total PIP >0.95, using individual level data for transformed protein levels and genotype dosages, iteratively increasing the maximum number of credible sets (*L*) starting from one. At each iteration step, variants were selected with the highest PIP from each CS, included in a joint regression model (with recruitment region, 11PCs, and array as covariates), and compared with the model from the previous iteration in R using anova(), until no additional CSs were found or the current iteration showed no improvement in model fit (P>5x10^-8^). The variant with the largest PIP in each Independent CS was classed as its lead variant.

### Comparison with UKB-PPP pQTLs

LD between CKB and UKB-PPP pQTLs was assessed using each of the 72K unrelated CKB LD reference and 350K unrelated UKB participants to determine the maximum r^2^ in either ancestry between each CKB CS lead variant and all corresponding UKB-PPP CS lead variants at the same locus; for assessment of D’ only the UKB reference was considered. CS overlap between a CKB CS and UKB-PPP pQTLs was assessed by scoring the presence of each CKB CS variant in all corresponding CSs reported at that locus for the UKB-PPP discovery dataset^21^. Cross-ancestry fine- mapping used SuSIEx^36^, with association signals categorised as being present in both ancestries where both CKB-specific and UKB-specific causal probabilities were >0.8. CKB CSs for which all three methods suggested no overlap with UKB signals were considered to be EAS specific.

### Functional annotation of pQTLs

Variant annotation of pQTLs was conducted according to Ensembl VEP 107^69^. We selected the most severe annotation for all variants across all available transcripts using BCFtools ‘split-vep -s worst’^70^. All variants within fine-mapped CSs were categorised according to their functional impact and location with respect to coding and transcript sequences, and according to whether they affected the gene encoding the target (i.e. assayed) protein, a gene for a different protein within the *cis* region, or at a *trans*-pQTL. Variants that had functional annotation as splice acceptor/donor, start lost, stop gained/lost, frameshift, missense, inframe insertion/deletion, and protein altering variant were considered to be PAVs.

### Phenome-wide pQTL to trait associations

GWAS Catalog v1.0.2 (accessed on 12/08/2024)^37^ was used to search for associations with the sentinel variants for each CKB *cis*-pQTL, or their proxies identified from the CKB 72K LD reference using the PLINK v1.9 `--ld-snp-list` command with an r^2^ threshold of 0.8 and LD-window of 500Kbp. For each trait or disease with associations derived from EAS-only GWAS, a single association was selected in the following order of priority: (i) sentinel variant; (ii) highest r^2^ proxy; (iii) smaller P-value for association with the outcome. Where necessary, mismatched variant coordinates were resolved using the R package biomart^71^. Associations that involved interaction terms were excluded. A similar search was separately conducted of multi-ancestry meta-analyses which included data from EAS-only studies.

Estimates of the magnitude of the association between protein level and disease outcome used the Wald ratio method; the change in the outcome due to a genetically-predicted 1SD increase in plasma protein levels was derived as the ratio of effect size for the variant-outcome association to that for the variant-protein association^72,73^. Outcome effect sizes reported as odds ratio (OR) were first natural log-transformed to convert to a beta estimate. Missing effect alleles for disease associations were identified by inspection of the original source publication (**Supplementary Table 16**).

### Colocalisation

Publicly-accessible genome-wide summary statistics for EAS-specific outcomes were retrieved either from GWAS Catalog or from Biobank Japan^40^ or National Biobank of Korea (KoGES)^41^ web resources (**Supplementary Table 17**) and, where necessary, converted to build GRCh38 using UCSC-liftOver^74^. Together with CKB summary statistics for the relevant protein, shared genetic variants within a genomic locus (as defined by LD clumping) were evaluated using the R package coloc^75^ to assess support for hypotheses: H0, implying no association with either trait; H1, denoting association with trait 1 but not trait 2; H2, indicating association with trait 2 but not trait 1; H3, suggesting association with both trait 1 and trait 2 as two separate association signals; and H4, indicating a shared association with both trait 1 and trait 2. We further analysed protein-trait pairs not reaching either PP.H3>0.8 or PP.H4>0.8 using the R package susieR^76,77^ which employs a fine-mapping framework to identify shared genetic signals when there are multiple casual SNPs. Colocalisation was performed using default variant priors.

### Drug targets

Existing drugs targeting the proteins with identified disease associations, were identified by searching publicly-available drug databases: Therapeutic Target Database^42^ (https://db.idrblab.net/ttd/, versionupdate: 10/01/24), Drug Bank^43^ (https://go.drugbank.com/, version 5.1.13), and Open Targets^44^ (https://www.opentargets.org/, release 24.09).

