## Supplementary Information and Figures for "Ancestry diversity in the genetic determinants of the human plasma proteome enhances identification of potential drug targets"

Members of the China Kadoorie Biobank Collaborative Group 2

Supplementary Figure 1. Distributions of measured protein levels (in NPX units) for multi-modal proteins. 3

Supplementary Figure 2. Comparison of sentinel variants from CKB *cis*-pQTLs with corresponding data from the Japan COVID-19 Task Force. 4

Supplementary Figure 3. Comparison of *cis*-pQTLs sentinel variants identified by the Japan COVID-19 Task Force. with corresponding data from CKB. 4

Supplementary Figure 4. Comparison of estimates for polygenic heritability using GCTA with estimates derived as $\Sigma(2pq\beta^2)$. 5

Supplementary Figure 5. Comparison of estimated heritability in CKB and UKB-PPP. 5

Supplementary Figure 6. Comparison of *cis*-pQTL variant effect sizes between CKB and UKB-PPP studies. 6

Supplementary Figure 7. Distribution of minor allele frequencies for lead variants from CKB credible sets. 6

Supplementary Figure 8. Distribution of the sum of posterior inclusion probabilities (PIP) for the variants in each CKB credible set which overlap with a corresponding UKB-PPP credible set. 7

Supplementary Figure 9. Comparison of LD and credible set overlap approaches for assessing signal ancestry specificity. 7

Supplementary Figure 10. Comparison of effect sizes for sentinel pQTL variants from the discovery GWAS with effect sizes from stratified GWAS. 8

Supplementary Figure 11. Comparison of-$\log_{10}$P values for sentinel pQTL variants from the discovery GWAS and meta-analysed case and subcohort GWAS. 8

**Members of the China Kadoorie Biobank Collaborative Group**

**International Steering Committee:** Junshi Chen, Zhengming Chen (PI), Robert Clarke, Rory Collins, Yu Guo, Liming Li (PI), Chen Wang, Jun Lv, Richard Peto, Robin Walters.

**International Co-ordinating Centre, Oxford:** Daniel Avery, Maxim Barnard, Derrick Bennett, Ruth Boxall, Ka Hung Chan, Yiping Chen, Zhengming Chen, Charlotte Clarke, Johnathan Clarke; Robert Clarke, Huaidong Du, Ahmed Edris, Hannah Fry, Simon Gilbert, Prapthi Harish, Pek Kei Im, Andri Iona, Maria Kakkoura, Christiana Kartsonaki, Kshitij Kolhe, Hubert Lam, Kuang Lin, James Liu, Mohsen Mazidi, Iona Millwood, Sam Morris, Qunhua Nie, Alfred Pozarickij, Maryam Rahmati, Paul Ryder, Becky Stevens, Iain Turnbull, Dan Valle Schmidt, Robin Walters, Baihan Wang, Lin Wang, Neil Wright, Ling Yang, Xiaoming Yang, Pang Yao.

**National Co-ordinating Centre, Beijing:** Jun Lv, Canqing Yu, Dianjianyi Sun, Yuanjie Pang, Can Hou, Qingmei Xia, Chao Liu, Pei Pei, Lang Pan, Xiao Han, Honglu Bian, Xinxin Chen.

**Regional Co-ordinating Centres:**

**Qingdao CDC:** Zengchang Pang, Ruqin Gao, Shanpeng Li, Haiping Duan, Shaojie Wang, Yongmei Liu, Ranran Du, Liang Cheng, Xiaocao Tian, Hua Zhang. **Licang CDC:** Dan Hu, Xiaoyan Zheng, Yujie Wang. **Heilongjiang Provincial CDC:** Wei Sun, Shichun Yan, Xiaoming Cui. **Nangang CDC:** Chi Wang, Zhenyuan Wu, Lishun Zhai, Zhaoxi Pang, Shiwen Dong. **Hainan Provincial CDC:** Huiming Luo, Jinyan Chen, Bin He, Dingwei Sun, Xingren Wang, Tingting Ou. **Meilan CDC:** Xiangyang Zheng, Dewei Zheng, Shuai Yang, Yilei Li, Lihui Li, Xingjiao Chen. **Jiangsu Provincial CDC:** Jinyi Zhou, Ran Tao, Jian Su, Xikang Fan, Zongming Cheng, Yuxiao Huang. **Suzhou CDC:** Yan Lu, Yujie Hua, Li Xing, Shuxian Wang, Jianrong Jin, Juping Ma, Jinchao Liu, Kaifei Zhu, Hongfu Ren, Xingfeng Shen. **Guangxi Provincial CDC:** Ge Zhong, Wei Mao, Zhenzhen Lu, Ling He. **Liuzhou CDC:** Lifang Zhou, Changping Xie, Jian Lan, Tingping Zhu, Jinxue Tan, Liuping Wei, Liyuan Zhou, Sisi Wang. **Sichuan Provincial CDC:** Xianping Wu, Ningmei Zhang, Xiaofang Chen, Xiaoyu Chang, Zhuo Wang, Yujin He. **Pengzhou CDC:** Mingqiang Yuan, Xia Wu, Xiaofang Chen, Zhaodong Wang, Qiang Sun, Yang Lin. **Gansu Provincial CDC:** Faqing Chen, Xiaolan Ren, Lijun Chang, Feiming Zhong. **Maiji CDC:** Jianjun Feng, Weijie Hu, Xiaofang Zhang, Yalin Chen, Fei Wang, Jun Wang. **Henan Provincial CDC:** Linqi Diao, Wanshen Guo, Zhiwei Han, Dongyang Zhao, Dengjun Zhu, Kai Kang, Shixian Feng, Huizi Tian, Yali Yan, Bing Han, Li Gao, Shaofang Li, Huafei Feng, Wei Tang. **Huixian CDC:** Xiaolin Li, Huarong Sun, Xiaocong Zhao, Ying Li, Chen Hu, Pan He, Xukui Zhang, Yuanyuan Jin, Hesheng Zhang. **Zhejiang Provincial CDC:** Min Yu, Ruying Hu, Hao Wang, Weiwei Gong, Jieming Zhong, Meng Wang, Chunxiao Xu, Keqing Gong. **Tongxiang CDC**: Hao Xu, Yuan Cao, Kaixu Xie, Lingli Chen, Xiaomei Tu,Chen Chen. **Hunan Provincial CDC:** Xiaojun Li, Li Yin, Huilin Liu, Yuan Liu, Yi Liu, Lei Yin, Xian Xie, Jing Wang. **Liuyang CDC:** Bo Xiao, Pingsheng Lou, Yuan Peng, Libo Zhang, Chan Qu, Qili Jiang, Yanling Chen, Yan Zhao.

### Supplementary Figures

**Supplementary Figure 1.** Distributions of measured protein levels (in NPX units) for multi-modal proteins.

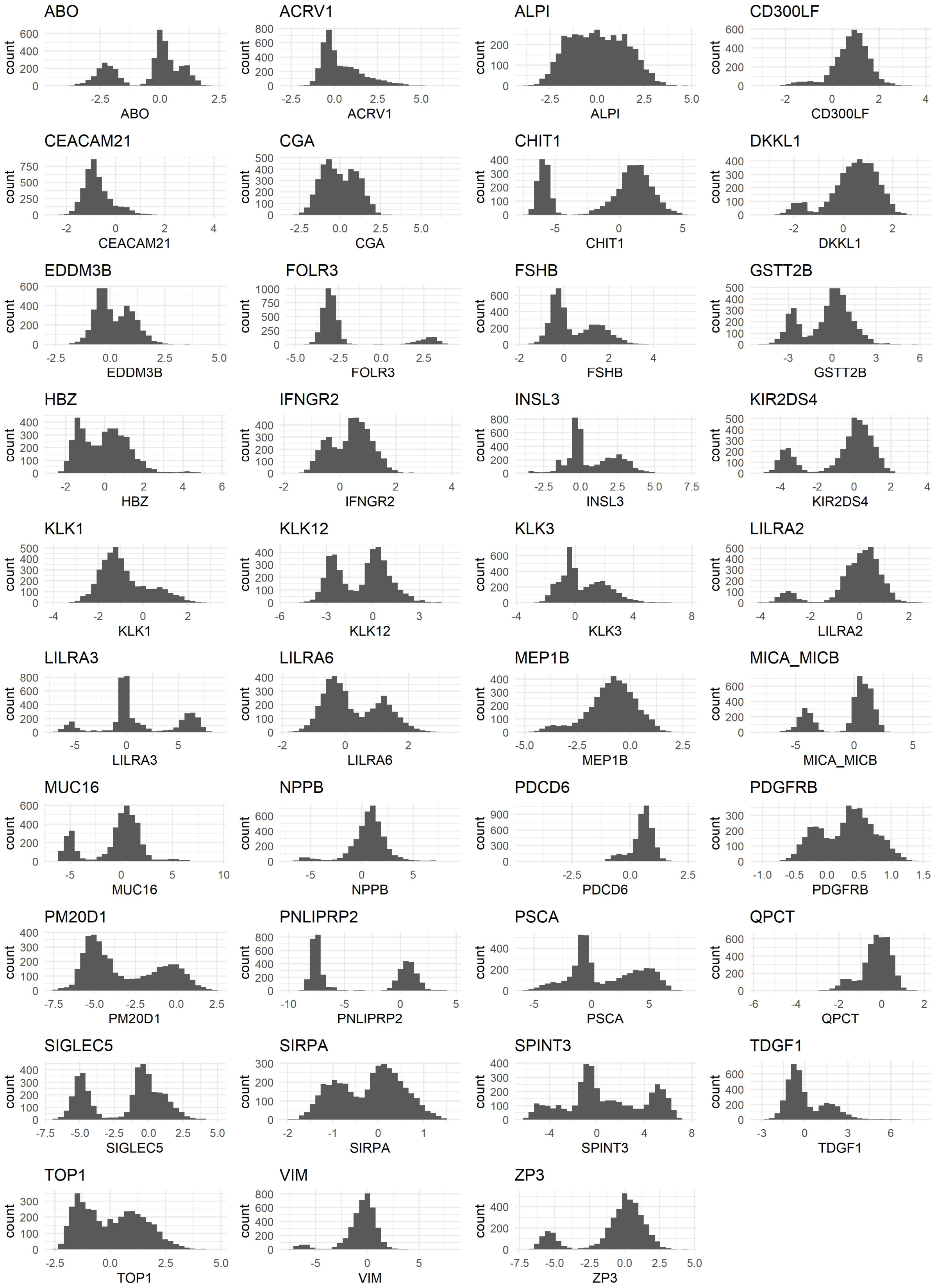

**Supplementary Figure 2**. Comparison of sentinel variants from CKB *cis*-pQTLs with corresponding data from the Japan COVID-19 Task Force: (A) effect allele frequency; (B) effect size. Where data were not available in the Japanese GWAS*,* the best available proxy from the CKB GWAS.

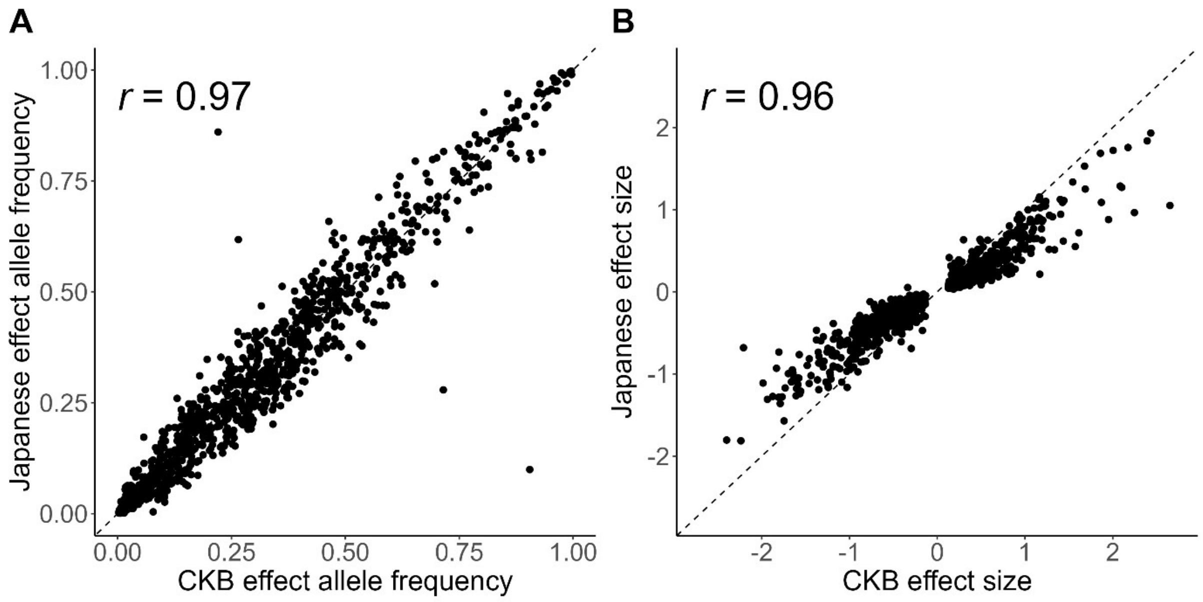


**Supplementary Figure 3.** Comparison of *cis*-pQTLs sentinel variants identified by the Japan COVID-19 Task Force. with corresponding data from CKB: (a) effect allele frequency; (B) effect size. Where variants were not available in the CKB GWAS the best available proxy from the discovery GWAS was used.

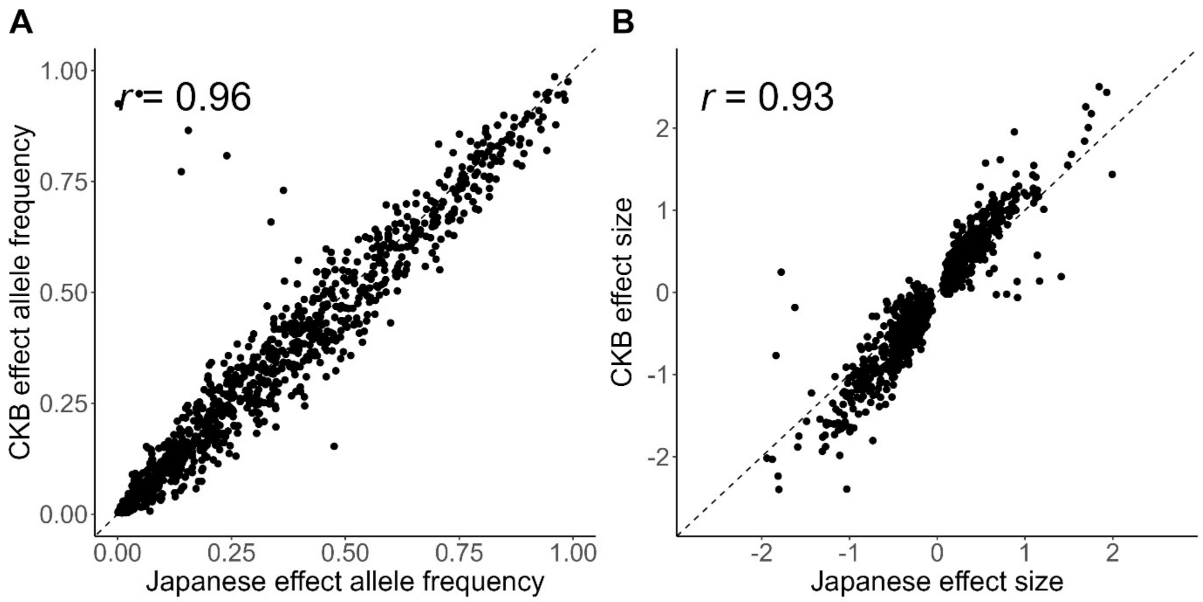

**Supplementary Figure 4.** Comparison of estimates for polygenic heritability using GCTA with estimates derived as $\Sigma(2pq\beta^2)$ for conditionally-independent variant associations: (A) total, (B) *trans*, (C) *cis* heritability.

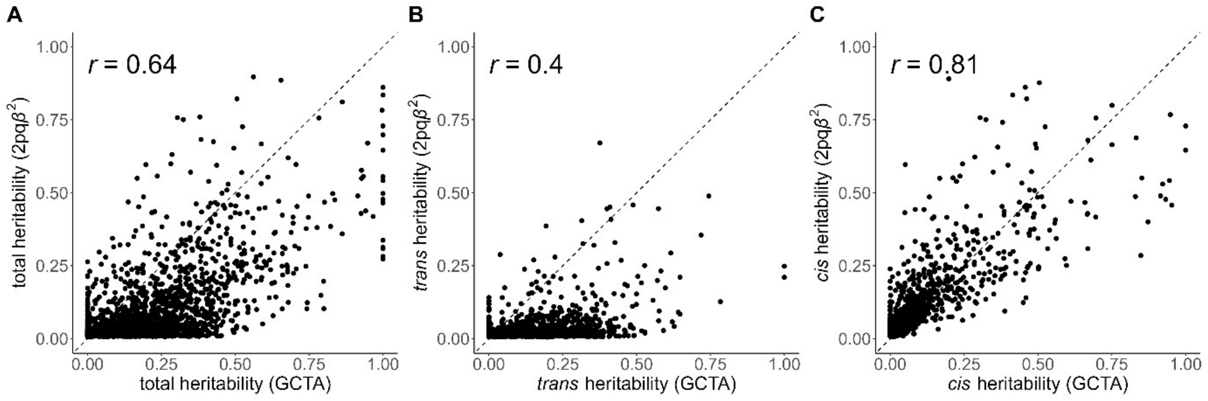


**Supplementary Figure 5.** Comparison of estimated heritability in CKB and UKB-PPP: (A) total polygenic heritability, (B) *cis* heritability.

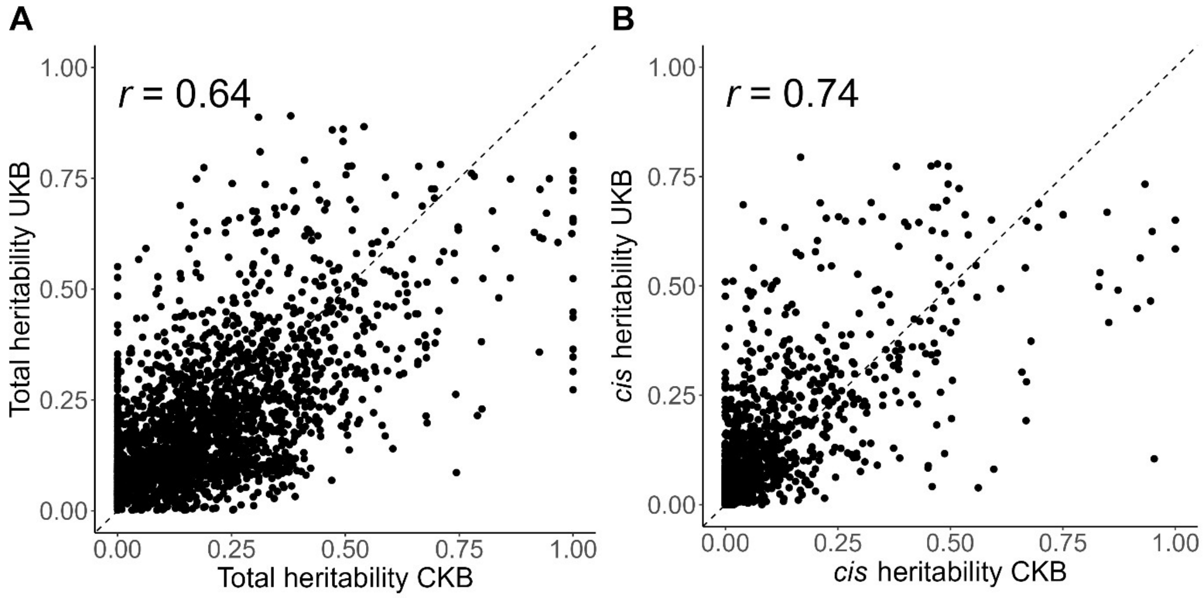

**Supplementary Figure 6.** Comparison of *cis*-pQTL variant effect sizes between CKB and UKB-PPP studies: (A) CKB sentinel variants; (B) UKB-PPP sentinel variants. Variants with data not present in the other study are not shown.

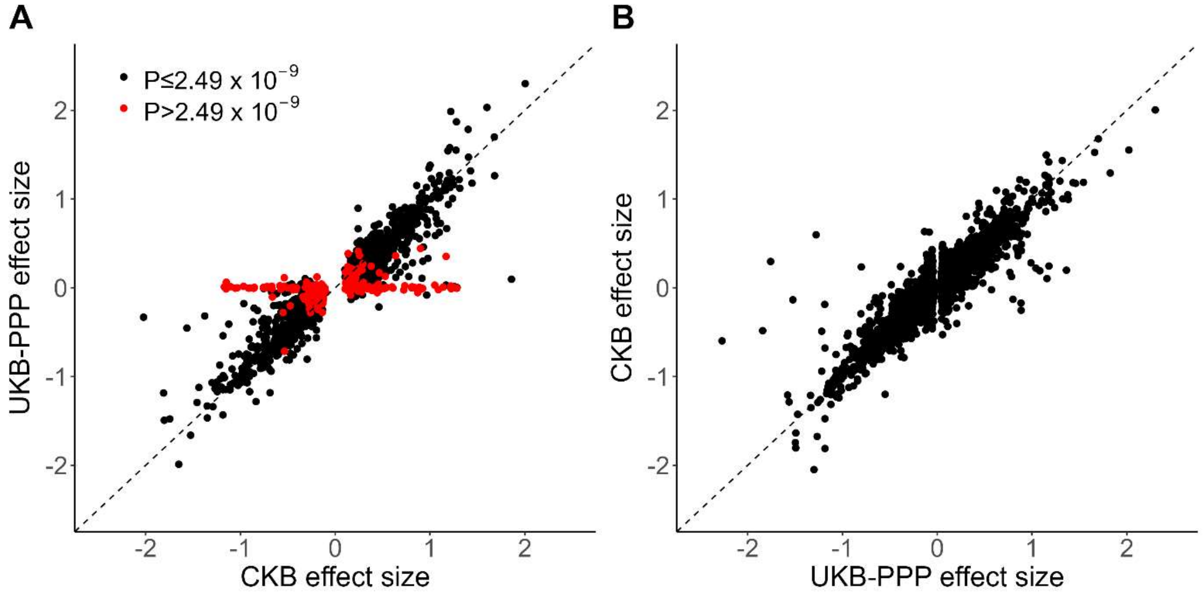


**Supplementary Figure 7.** Distribution of minor allele frequencies for lead variants from CKB credible sets, according to the maximum LD between UKB and UKB-PPP lead variants. (A) in non-Finnish Europeans according to gnomAD; (B) in CKB.

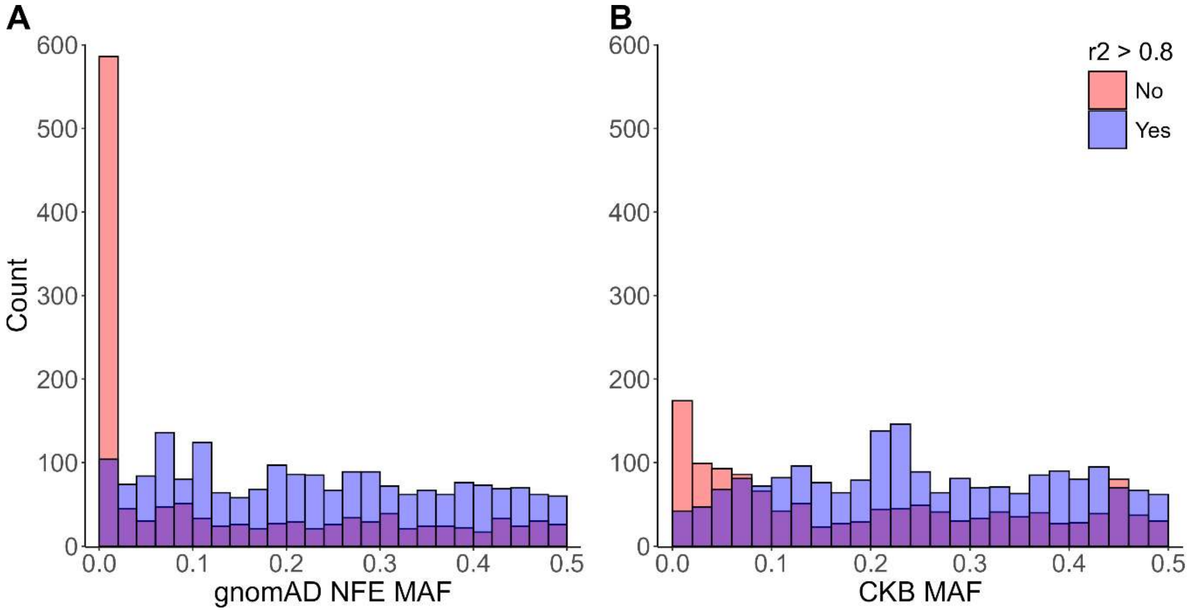

**Supplementary Figure 8.** Distribution of the sum of posterior inclusion probabilities (PIP) for the variants in each CKB credible set which overlap with a corresponding UKB-PPP credible set.

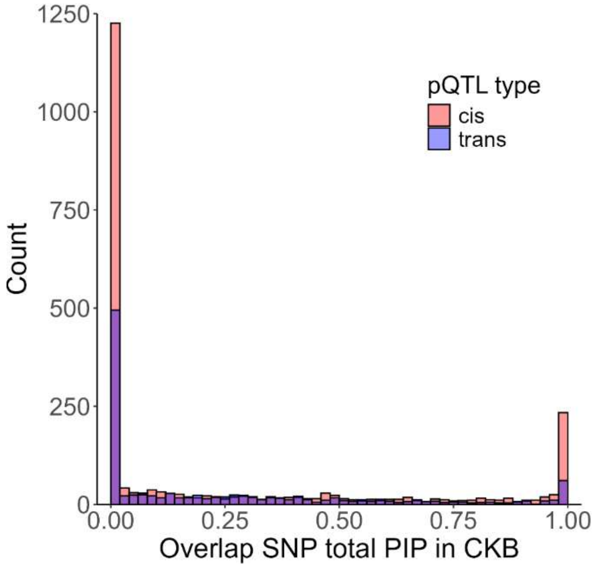


**Supplementary Figure 9. Comparison of LD and credible set overlap approaches for assessing signal ancestry specificity.** (A) Distribution of the maximum $r^2$ between lead variants for CKB and UKB-PPP pQTL credible sets, according to whether CKB credible sets have any overlap with the corresponding UKB-PPP credible sets. (B) Distribution of the sum of posterior inclusion probabilities (PIP) for the variants in each CKB credible set which overlap with a corresponding UKB-PPP credible sets, according to whether the lead variant is in LD with a UKB-PPP lead variant.

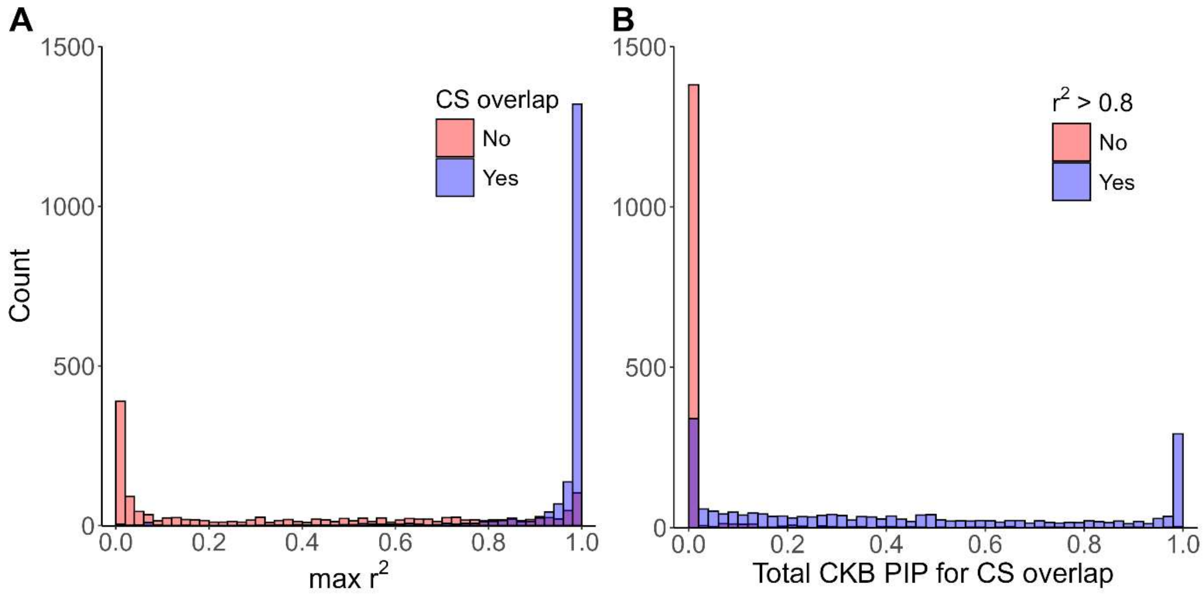

**Supplementary Figure 10.** Comparison of effect sizes for sentinel pQTL variants from the discovery GWAS with effect sizes from stratified GWAS in (A) subcohort only and (B) cases only.

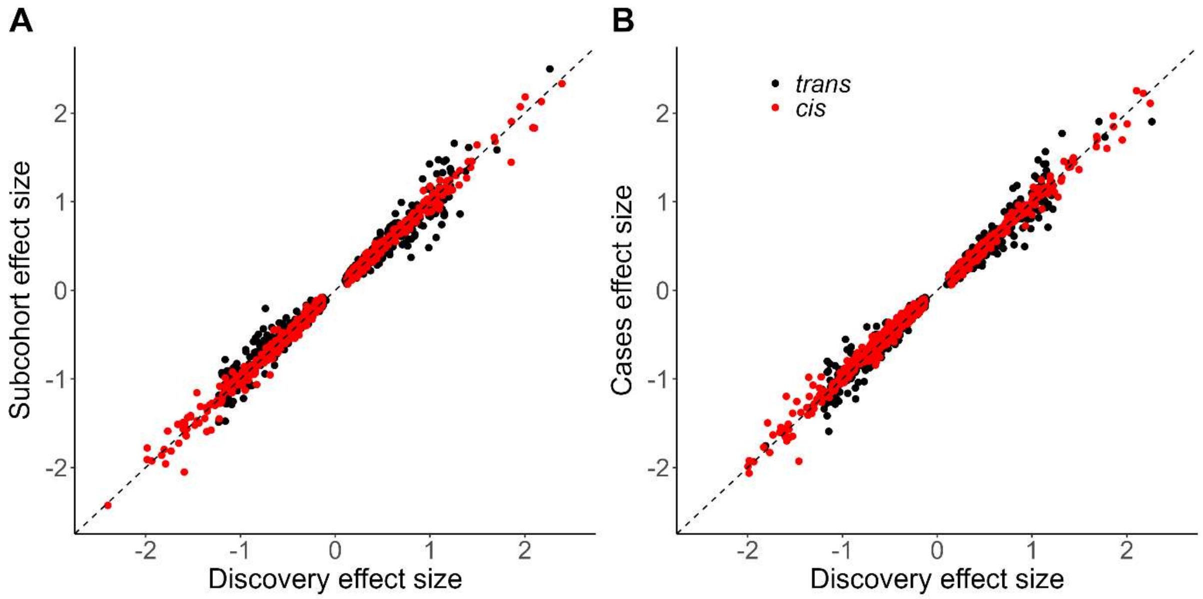


**Supplementary Figure 11**. Comparison of-$\log_{10}P$ values for sentinel pQTL variants from the discovery GWAS and meta-analysed case and subcohort GWAS.

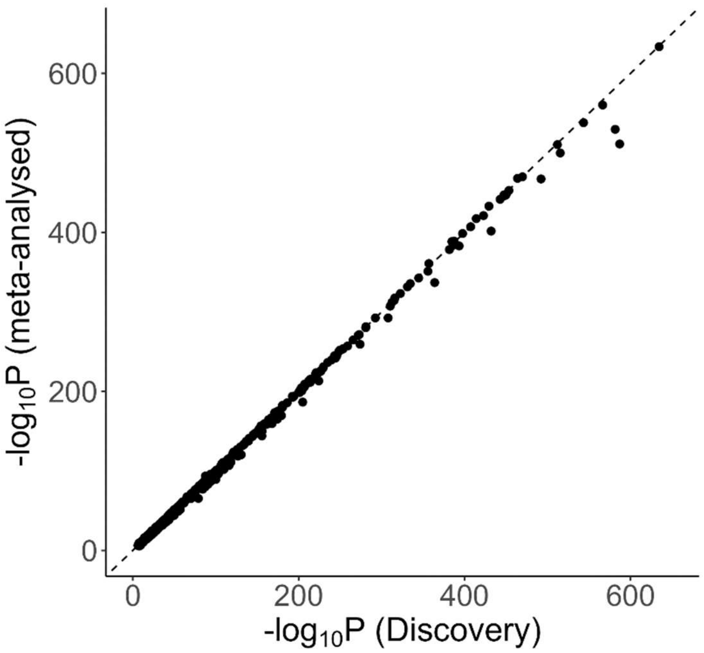
